# An interpretable whole-blood transcriptomic model for Parkinson’s disease identification and molecular profiling

**DOI:** 10.64898/2026.10.19.26365876

**Authors:** Yang Li, Xiya Yu, Changgeng Peng

## Abstract

**Background:** Sporadic Parkinson’s disease (PD) lacks a molecular taxonomy that stratifies patients by dominant molecular drivers, limiting enrollment for disease-modifying trials, especially anti-inflammatory agents targeting NLRP3/NF-κB.

**Methods:** We developed ORIGIN-PD, an interpretable whole-blood transcriptomic classifier comprising 34 gene-level features and a composite low-expression score, with gene-level contributions organized into eight prespecified functional modules: midbrain dopaminergic (mDA) neuron identity/development, pathway-related, oxidative/mitochondrial, proteostasis, inflammation/ immunity, microglia, ncRNA, and other. Using LASSO regression on stringently quality-controlled whole-blood RNA-seq data from the Parkinson’s Progression Markers Initiative (PPMI) (Training Set: 204 PD and 92 controls; Locked Test Set: 51 PD and 23 controls), we assigned each patient a module-contribution profile. Module dominance was defined as >30% of total score.

**Results:** ORIGIN-PD discriminated PD from controls with high accuracy (Locked Test Set AUC 0.944, sensitivity 0.843, specificity 0.913). Among 255 sporadic PD patients, 49.4% exhibited a dominant module-contribution pattern—most frequently mDA neuron identity/development (22.0%), followed by pathway-related (11.8%), microglia/glial (7.5%), and inflammation/immunity (3.5%). Among the 50.6% with multi-module contributions, mDA neuron identity/development ranked in the top three in 62.8% of patients versus 35.7% for inflammation/immunity, indicating broader involvement of mDA neuron identity genes than inflammatory genes. The model showed high specificity against Alzheimer’s disease, ALS, and inflammatory controls.

**Conclusions:** ORIGIN-PD enables rational enrichment for NLRP3/NF-κB inhibitor trials by identifying patients with inflammatory involvement. In addition, the predominant contribution of the mDA neuron identity/development module, centred on *WNT1*, *MSX1*, *NPM1*, and *LMX1B*, identifies a larger patient subgroup with altered expression of mDA neuron identity module genes, suggesting that ORIGIN-PD may also be useful for patient enrichment in future trials of Wnt/β-catenin pathway agonists. This dual-purpose framework provides a transparent, interpretable basis for pathogenic subtyping and precision enrichment in sporadic PD, with the aim of guiding disease-modifying therapy.

## Background

Parkinson’s disease (PD) is a clinically and biologically heterogeneous neurodegenerative disorder characterized by progressive motor and non-motor manifestations[1–3]. Despite advances in neuroimaging and molecular biomarker research, the diagnosis of PD remains primarily based on clinical history and neurological examination[4, 5]. Moreover, substantial inter-individual variability exists in disease presentation, progression and underlying molecular pathology[2, 6], suggesting that PD may arise from partially distinct biological processes across individuals. Accessible molecular tools capable of complementing clinical assessment while capturing this biological heterogeneity are therefore needed.

We recently found that Nucleophosmin 1 (NPM1/B23) quantitatively regulates the number and survival of substantia nigra pars compacta (SNc) dopaminergic (DA) neurons through WNT1 signaling pathway in mice (our unpublished data). These findings, together with those from other laboratories[7–11], suggest that NPM1/WNT1 may be therapeutic targets for sporadic PD. In addition to the intrinsic resistance conferred by the gene expression profile of SNc DA neurons, external stresses such as inflammation also critically contribute to the death of these neurons through NLRP3 (NLR family pyrin domain containing 3) inflammasome activation and/or NF-κB signaling[12–14]. These pathways are key inflammatory mediators in PD progression [15, 16]. Accordingly, anti-inflammatory agents targeting NLRP3 or NF-κB-related signaling have entered early-phase clinical evaluation in PD[17, 18]. These disease-modifying therapies could advance PD treatment. However, it remains unknown whether all sporadic PD patients are affected by inflammatory or NPM1/WNT1 signaling pathways. If not, an accessible method is required to stratify PD patients for specific targeted therapies.

Peripheral blood represents an attractive source for biomarker development because of its accessibility, scalability and potential to reflect systemic molecular alterations associated with PD[3, 19–22]. Transcriptomic profiling has revealed widespread changes in peripheral blood involving immune regulation, metabolism, proteostasis, mitochondrial function, and other biological processes implicated in PD pathogenesis[19, 20, 23]. Machine-learning approaches provide an opportunity to integrate these high-dimensional molecular signals into clinically informative signatures[24, 25]. However, several challenges remain, including the robustness of feature selection, false-positive classification, transferability across independent cohorts and biological interpretation of model predictions. In particular, models optimized solely for overall discrimination may provide limited insight into the molecular processes contributing to individual predictions[25, 26].

Although whole-blood transcriptomes cannot directly mirror SNc DA neuron-specific expression, they capture systemic molecular programs that may partially reflect the same pathogenic axes. We therefore sought to develop a module-based scoring framework as a biologically interpretable proxy. We hypothesized that the intrinsic expression profile of SNc DA neurons, including genes involved in their development and maintenance, and external factors such as inflammation, oxidative stress, and environmental exposures, contribute differently to PD across individuals, and that these contributions could be captured using whole-blood transcriptomic profiles. To this end, we therefore predefined eight PD-relevant functional modules and integrated data-driven transcriptomic features with biologically informed candidates to develop ORIGIN-PD (Octa-module RNA-seq Investigation for Genesis and Initiation of Neurodegenerative Parkinson’s Disease), an interpretable model for PD identification and patient stratification. The resulting contribution scores for individual patients may help guide the selection of the most suitable disease-modifying therapy.

## Materials and methods

### 1. Study design, participants and transcriptomic data

This study developed ORIGIN-PD as a cross-sectional whole-blood transcriptomic classifier for discriminating PD from healthy control (HC). The primary dataset was obtained from the PPMI. Baseline whole-blood RNA-sequencing samples were matched to participant-level clinical metadata, and technical, pooled, or otherwise non-eligible samples that did not represent participant-level baseline observations were excluded.

RNA-sequencing sample quality was assessed using RNA integrity number (RIN), uniquely mapped reads, the number of detected genes, the intergenic-read ratio, and the proportion of usable sequencing bases. Samples were required to meet all prespecified quality-control criteria: RIN ≥7, uniquely mapped reads ≥60%, at least 30,000 detected genes, an intergenic-read ratio ≤13%, and usable sequencing bases ≥30%. After sample-level quality control and restriction to baseline PD and HC participants, the final dataset from PPMI cohort comprised 370 participants, including 255 with PD and 115 HC.

Dataset partitioning was performed at the participant level so that repeated observations from the same participant could not cross analysis sets. A fixed diagnosis-stratified split was generated with random seed 20260419. The Training Set comprised 296 participants (204 PD and 92 HC), and the Locked Test Set comprised 74 participants (51 PD and 23 HC). The Locked Test Set was reserved as an internal independent test set and used exclusively for final evaluation of model performance.

### 2. RNA-sequencing preprocessing and transcriptomic feature construction

#### 2.1 RNA-sequencing preprocessing

Gene-level raw feature counts output was used as the transcriptomic input. Ensembl version suffixes were removed before feature alignment, and duplicate Ensembl identifiers were collapsed by taking the mean expression value across duplicate rows. Genes were then filtered according to expression abundance and prevalence across all RNA-seq samples passing sample-level quality control, retaining genes with CPM ≥1 in at least 10% of the sample. This reduced the transcriptomic feature space from 58,735 unique genes to 21,785 genes. Normalization factors were estimated using the trimmed mean of M-values (TMM) method implemented in edgeR, after which expression values were transformed to log2 counts per million (log2CPM) with a prior count of 1.

#### 2.2 Predefined biological framework

Before model training, eight PD-relevant functional modules were defined to represent distinct biological processes potentially contributing to disease heterogeneity: mDA neuron identity/development, inflammation/immunity, oxidative/mitochondrial function, non-coding RNA regulation, proteostasis/clearance, pathway-related signaling, microglial/glial responses, and other PD-related processes. For each module, biologically informed candidate genes were prespecified on the basis of prior mechanistic and literature evidence (**Table S1**). These module-associated candidates were incorporated alongside data-driven transcriptomic features during model development, while final feature retention and weighting were determined by the modeling procedure rather than by module membership. Several prespecified candidates were retained in ORIGIN-PD, allowing the same biological framework to be used subsequently for participant-level contribution analysis.

#### 2.3 Differential-expression gene (DEG) analysis and DEG-derived feature selection

DEG analysis between PD and HC participants was performed using limma, with diagnosis, sex, sequencing plate, age, and usable sequencing bases included as model terms, followed by empirical Bayes moderation with trend and robust estimation. Genes with nominal *P* < 0.05 and absolute log2 fold change > 0.1 were retained, yielding 1,044 candidates. These were ranked by ascending nominal *P* value, descending absolute log2 fold change, and ascending adjusted *P* value, and the top 100 unique genes were carried forward.

Feature stability was evaluated across 50 diagnosis-stratified resampling iterations using least absolute shrinkage and selection operator (LASSO) logistic regression. Genes showing consistent selection and coefficient direction were retained, followed by incremental performance evaluation and backward elimination based on cross-validated performance within the Training Set. Features were removed when their exclusion preserved specificity and decreased balanced accuracy and ROC-AUC by no more than 0.005 each. This procedure yielded the final 35-gene DEG-derived panel (**Table S2**).

#### 2.4 Construction of the low-expression transcriptomic signature

Low-abundance transcripts were evaluated as a separate feature-construction branch because coordinated variation across individually sparse transcripts can carry information that is not well represented by single low-expression genes. Within the Training Set, low-expression candidates were defined as transcripts with CPM >0.1 in at least 10% of samples but CPM >1 in fewer than 10% of samples, yielding 14,994 candidate transcripts.

Candidate stability was assessed in 100 diagnosis-stratified subsamples, each containing 80% of Training Set participants. Differential-expression analysis within each subsample used the edgeR quasi-likelihood framework with diagnosis, sex, sequencing plate, age, and usable sequencing bases included as covariates. Genes were retained if they met the nominal significance and effect-size criteria (P<0.05 and |log_2_FC| >0.10) in at least 50% of subsamples, ≥80% directional consistency, and |median log_2_FC| ≥0.10, and were not flagged as outlier-prone. This yielded a 622-gene reference set comprising 228 PD-upregulated and 394 PD-downregulated genes.

For participant *i* and low-expression gene *g*, log2CPM expression was standardized using the Training Set mean and standard deviation:

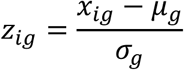

where xig denotes the log2CPM expression of gene g in participant i, and μg and σg denote the mean and standard deviation, respectively, of gene g in the Training Set. The same Training Set-derived *μ_g_* and *σ_g_* values were used for all subsequent datasets.

To improve the robustness of the low-expression signature, the 622 stable genes were further ranked according to a gene-level stability metric. Candidate pruning depths were evaluated by repeated cross-validation within the PPMI development cohort, supporting a pruning depth of 300 and leaving 322 active contributors. The final LowExpr_score was calculated as a weighted sum of standardized expression values across these active genes using the fixed direction-specific weights defined in the original signature; no post-pruning renormalization was applied. The composite score was calculated as:

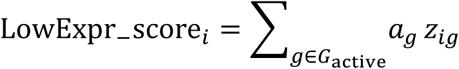

where *G_active_* denotes the 322 active low-expression genes and *a_g_* denotes the fixed direction-specific weight for gene. Only the 322 retained genes contributed to LowExpr_score.

#### 2.5 Investigator-specified PD/mDA-related candidate genes

Forty-three prespecified module-associated genes (**Table S1**) were evaluated jointly with the DEG-derived candidates in an initial biologically informed screening. However, none of these candidates met the predefined retention criteria, suggesting that the highly heterogeneous and correlated feature space may have introduced substantial redundancy and competition among signals, thereby limiting stable incremental contributions in the multivariable LASSO framework. Nine genes were therefore prespecified for biological evaluation on the basis of prior evidence linking them to PD-related biology, mDA identity/development, or DA neuronal maintenance: *NPM1, WNT1, CTNNB1, LEF1, BDNF, PITX3, NRP2, LMX1B,* and *MSX1*[8, 27–34]. The 35 DEG-derived gene-level candidates plus LowExpr_score constituted an existing 36-input transcriptomic set. Each of the nine investigator-specified genes was evaluated jointly with this 36-input set using 50 repeats of fivefold cross-validation in the Training Set.

Five genes met the implemented selection-frequency criterion of at least 60%: *NPM1, WNT1, BDNF, LMX1B,* and *MSX1*. The resulting candidate space therefore contained 35 DEG-derived genes, LowExpr_score, and five retained investigator-specified genes, for a total of 41 model inputs (40 gene-level candidates plus LowExpr_score). This 41-input candidate space was taken forward for Primary Model development.

### 3. Development and refinement of ORIGIN-PD

#### 3.1 Development of the Primary Model

The Primary Model was developed from the verified 41-input candidate space using LASSO logistic regression in the Training Set only[35]. Features were standardized, and binomial models were fitted with glmnet α=1[36]. Ten-fold cross-validation was used to select the regularization parameter by the one-standard-error criterion, with ROC-AUC as the cross-validation performance measure. The fixed cross-validation seed for the final Primary Model fit was 20260701.

The selected Primary Model retained 29 non-zero inputs, comprising 28 gene-level features (*MAP4K4, COL9A3, SRPK1, GSTZ1, CDK6, SOD2, WNT1, NRL, TOR1B, LMX1B, NRM, TMEM45B, OLAH, MSX1, CACNB3, SNHG11, BDNF, NPM1, CD300LD, LINC00623, WDR86-AS1, CD8B2, CEBPA-DT, PTGES3P1, lnc-AMDHD1-1, lnc-FBXO25-2, lnc-TBPL2-6* and *AC007298.1*) and LowExpr_score. Repeated fivefold cross-validation was additionally used during development to assess the robustness of the low-expression pruning strategy and the resulting classifier. Apparent Training Set performance from the final fit was kept distinct from these repeated cross-validated estimates.

#### 3.2 Candidate-space expansion and development of the Intermediate Model

To broaden biological coverage and assess whether complementary mechanistic signals not captured by the Primary Model candidate space could provide additional discriminatory information, we expanded the candidate space from the pre-Primary 36-input transcriptomic base, rather than the 29-input Primary Model. We added 59 genes, including biologically informed candidates from the prespecified eight-module framework (**Table S1**) and protein– protein interaction (PPI)-associated genes linked to features in the original 36-input set, yielding a 95-input candidate space. The identities, sources, and biological categories of these genes are summarized in **Table S3**.

The 95-input candidate space was evaluated by LASSO in the Training Set[35, 36]. We performed repeated nested cross-validation with 50 repetitions of fivefold outer cross-validation. Within each outer-training partition, the regularization parameter was selected by fivefold inner cross-validation using the one-standard-error rule, with ROC-AUC as the optimization metric[37]. A final Training Set fit used tenfold cross-validation. This procedure retained 35 non-zero inputs, comprising 34 gene-level features and LowExpr_score, and defined the Intermediate Model.

#### 3.3 Constrained refinement and freezing of ORIGIN-PD

A final constrained refinement was performed to evaluate whether *NPM1* could replace *HSPA8* while preserving the multivariable structure of the Intermediate Model, motivated by the strong correlation between their expression levels in both the Training Set and Locked Test Set. *HSPA8* was replaced by *NPM1*, while the coefficients of the remaining 33 gene-level features and LowExpr_score were held fixed. Only the model intercept and the *NPM1* coefficient were re-estimated by logistic regression in the Training Set.

The resulting classifier was finalized as ORIGIN-PD and comprised 35 model inputs: 34 gene-level features and LowExpr_score. For participant *i*, the linear predictor and corresponding PD probability were calculated as:

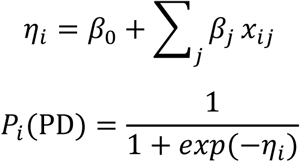

where *β_0_* denotes the model intercept, *β_j_* the coefficient of feature *j*, and *x_ij_* the value of feature *j* for participant *i*.

All model coefficients, feature definitions, normalization parameters, and LowExpr_score scaling parameters were fixed before downstream evaluation. The operating threshold of 0.69 was originally selected in the Training Set by maximizing balanced accuracy across candidate cut-offs applied to diagnosis-stratified 10-fold out-of-fold predictions and was subsequently retained unchanged for ORIGIN-PD. Probabilities ≥0.69 were classified as PD-like, whereas probabilities <0.69 were classified as HC-like. ORIGIN-PD was developed to distinguish PD from HC at the time of assessment and was not designed to predict future PD risk.

### 4. Internal model evaluation and biological interpretation

#### 4.1 Model performance and calibration

Model evaluation distinguished three quantities: apparent Training Set performance from a final fitted model, cross-validated Training Set performance used during feature selection or model comparison, and Locked Test Set performance obtained by applying a model without refitting. Apparent Training Set estimates were not described as cross-validated performance.

Discrimination was summarized by ROC-AUC and precision–recall AUC (PR-AUC). Probabilistic performance was assessed using the Brier score and binary log-loss[38]. At the fixed 0.69 threshold, sensitivity, specificity, accuracy, balanced accuracy, positive predictive value (PPV), negative predictive value (NPV), F1 score, and the full true-positive/true-negative/false-positive/false-negative confusion matrix were reported.

ROC-AUC confidence intervals were calculated using the DeLong method where applicable[39]. Threshold-based and other performance estimates were accompanied by percentile confidence intervals from outcome-stratified bootstrap resampling; 2,000 bootstrap replicates were used for the reported internal performance estimates and paired model comparisons. Calibration was evaluated using quantile-binned calibration curves together with calibration-in-the-large and calibration slope[40].

#### 4.2 Feature contribution and direction analysis

Feature-level interpretation used the frozen ORIGIN-PD coefficients. Because ORIGIN-PD is a linear logistic model, analytical SHAP values were calculated on the log-odds scale[41], using the Training Set mean of each model input as the common reference background:

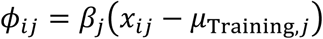

where *ϕ_ij_* denotes the contribution of feature *j* to participant *i*, *β_j_* is the corresponding model coefficient, *x_ij_* is the observed feature value, and *μ_Training,j_* is the mean value of feature *j* in the Training Set.

The same Training Set reference background was used to calculate SHAP values for participants in both the Training Set and Locked Test Set, and global feature importance was summarized by mean absolute SHAP value[41]. For visualization, SHAP values from the two sets were combined after calculation; no pooled reference background was recalculated, and the model itself was not refitted or redefined using the combined data.

To distinguish marginal expression differences from conditional multivariable contributions, PD–HC differences for each of the 34 gene-level features were quantified using Hedges’ g separately in the Training Set and Locked Test Set[42]. Directional consistency was assessed by comparing three quantities: the marginal PD–HC expression direction, the direction of association from univariate logistic regression, and the sign of the corresponding ORIGIN-PD multivariable coefficient. Reproducibility of marginal effects between the two sets was further evaluated by directional concordance and Spearman correlation of gene-level Hedges’ g estimates. Because Hedges’ g reflects the marginal standardized PD–HC difference for an individual feature, whereas a multivariable coefficient reflects its conditional contribution in the context of the remaining model features, sign reversals may occur when correlated features are modeled jointly[43].

#### 4.3 Functional-module and individual contribution analysis

For biological organization, the 34 ORIGIN-PD gene-level features were assigned a priori to eight functional categories: pathway-related, inflammation/immunity, oxidative/mitochondrial, non-coding RNA, proteostasis/clearance, mDA neuron identity, microglia/glial, and other. These categories were prespecified for model interpretation and were not derived from post hoc pathway-enrichment analysis. The 322 active low-expression genes contributing to LowExpr_score were annotated separately for descriptive composition; they remained components of a single composite model input.

Participant-specific signed module contributions were calculated on the same Training Set-centered log-odds scale used for analytical SHAP. For participant *i* and module *m*:

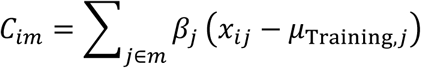

where *C_im_* denotes the signed contribution of module *m* to participant *i*, summed across all gene-level features assigned to that module.

Modules were ranked for each participant by the absolute magnitude of their signed contribution. LowExpr_score and the intercept were retained in the full ORIGIN-PD probability but were excluded from module ranking and module-share calculations. The primary module share was defined as the absolute contribution of the highest-ranked module divided by the sum of absolute contributions across all eight gene-level modules.

A strong contribution pattern was defined by two criteria: the dominant module accounted for at least 30% of the total absolute module contribution, and its absolute contribution was at least 25% greater than that of the second-ranked module:

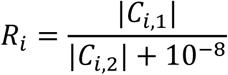

where *C_i,1_* and *C_i,2_* denote the largest and second-largest absolute module contributions for participant *i*, respectively. Participants were classified as having a strong contribution pattern when *R_i_* ≥ 1.25 and the dominant-module share was at least 30%; all others were classified as having mixed contribution patterns for descriptive analyses.

Module ranks were assigned independently for each participant according to the absolute magnitude of module contributions. Top-three summaries were calculated both across all PD participants and separately within the multimodule-contribution (MC) subgroup. For MC participants, first-, second-, and third-ranked modules were summarized directly, whereas monomodul-dominant (MD) participants were characterized primarily by their dominant (rank-1) module. MD participants in which a given module ranked second or third were retained when calculating top-three frequencies across the full PD cohort.

Representative individual contribution profiles were selected to illustrate the range of model-derived molecular patterns. These profiles and module-rank summaries describe contributions within the classifier and were not interpreted as biological subtypes, pathway activation states, cell-type abundance estimates, or causal disease mechanisms.

### 5. External evaluation and disease-control stress testing

Model normalization was performed in R using edgeR (version 4.4.0) with a Training Set-derived reference, with at least 80% reference-gene coverage required for reference-assisted normalization. All features required by the finalized classifier, including those used to construct LowExpr_score, were required to be available, and missing model inputs were not imputed. External datasets with incomplete feature coverage were analyzed only under a predefined sensitivity-analysis procedure described below.

#### 5.1 External PD/HC evaluation

GSE165082 was used as an external PD/HC evaluation cohort and comprised 26 participants (12 PD and 14 HC)[44]. External expression features were mapped to the ORIGIN-PD input space, normalized within the external dataset, and aligned to the PPMI Training Set control reference distribution using the implemented gene-wise mapping procedure. The ORIGIN-PD coefficients and the 0.69 threshold were then applied without feature selection, coefficient refitting, recalibration, or threshold optimization. Discrimination and threshold-based performance were summarized using the same applicable metrics used for internal evaluation.

#### 5.2 Neurological disease-control assessments

Non-PD neurological and neurodegenerative datasets were analyzed as cross-disease specificity stress tests rather than as PD sensitivity-validation cohorts. GSE277709 comprised 42 HC and 42 participants with motor neuron disease/amyotrophic lateral sclerosis and provided near-complete coverage, including 320 of the 322 active low-expression genes[45]. The ORIGIN-PD model was applied without refitting.

GSE270454 comprised 25 HC, 10 participants with mild cognitive impairment, and 10 participants with Alzheimer’s disease[46]. This dataset contained 31 of the 34 ORIGIN-PD gene-level features and 113 of the 322 active low-expression genes. Because feature coverage was incomplete, this analysis was designated an exploratory sensitivity assessment rather than a complete-input model projection. Under the predefined incomplete-feature sensitivity procedure, unavailable components were assigned zero contribution and the available-feature coverage was reported explicitly.

#### 5.3 Healthy and systemic disease transferability assessments

Additional datasets were used to assess transfer behavior in healthy or non-neurological disease settings. SoundLife contributed 358 analyzed observations, including 142 selected baseline-like observations and 216 other observations[47]. GSE178491 was used as an inflammatory-disease stress test[48] and GSE185263 as a sepsis/critical-illness stress test[49]. The analyzed GSE178491 dataset comprised 30 comparator observations and 135 inflammatory-disease observations, whereas GSE185263 comprised 44 HC and 348 sepsis observations.

### 6. Statistical analysis

Continuous variables were summarized as mean ± standard deviation or median [interquartile range], as appropriate, and categorical variables as counts and percentages. Standardized mean differences (SMDs) were used to assess baseline balance between the Training Set and Locked Test Set. All statistical tests were two-sided unless otherwise specified, with P < 0.05 considered nominally significant. Multiple-testing correction was applied where specified for high-dimensional analyses.

Wilcoxon rank-sum and Kruskal–Wallis tests were used for non-parametric group comparisons, as appropriate. Marginal PD–HC differences in gene expression were quantified using Hedges’ g[42], and Spearman rank correlation was used for association and cross-set reproducibility analyses. Confidence intervals for ROC-AUC were estimated using the DeLong method[39], whereas percentile bootstrap confidence intervals were used for threshold-based performance estimates and paired model comparisons. Model calibration was assessed using calibration curves, calibration-in-the-large, and calibration slope[40].

All analyses were performed in R version 4.4.3. RNA-sequencing normalization and log2CPM transformation were implemented using edgeR (version 4.4.0)[50], and differential-expression analysis was performed using limma[51]. Penalized logistic regression was implemented using glmnet (version 4.1-10)[36]. Additional package versions and computational details are provided in the reproducibility materials.

## Results

### Study cohort and transcriptomic feature construction for PD discrimination

We used a whole-blood RNA-sequencing dataset from the PPMI cohort to develop a PD discrimination model. The study workflow integrated PPMI cohort construction, complementary feature engineering, sequential model development, and evaluation of the ORIGIN-PD model (**Figure 1**). After matching samples to clinical metadata, only eligible baseline whole-blood RNA-sequencing samples from PD and HC were retained; technical and pooled samples were excluded. Sample-level RNA-sequencing quality control required an RIN ≥7, ≥60% uniquely mapped reads, ≥30,000 detected genes, an intergenic ratio ≤13%, and ≥30% usable bases. The final PPMI dataset comprised 370 participants, including 255 PD and 115 HC. Participants were assigned to a Training Set of 296 individuals (204 PD and 92 HC) and a Locked Test Set of 74 individuals (51 PD and 23 HC). Baseline demographic, clinical, and sequencing characteristics were well balanced between the Training Set and Locked Test Set, with standardized mean differences indicating no substantial imbalance across the principal variables examined (**Table 1**). All feature-selection and model-development procedures were performed within the Training Set. The Locked Test Set was not used for coefficient estimation, lambda selection, or threshold fitting.

**Fig. 1.**
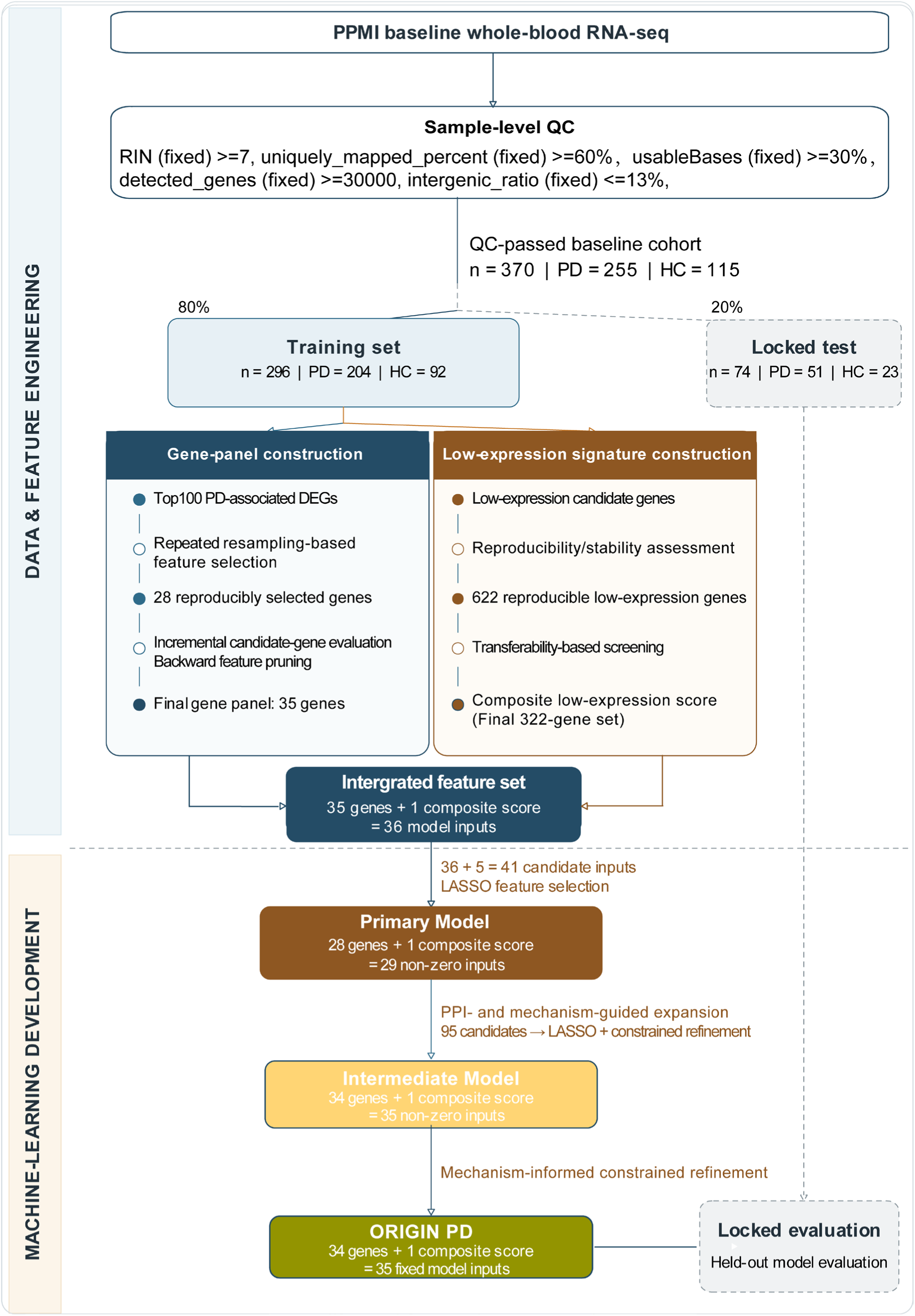
Study design and model-development workflow for ORIGIN-PD. Baseline whole-blood RNA-sequencing data from the Parkinson’s Progression Markers Initiative (PPMI) were evaluated for sample quality, including RNA integrity, mapping, gene detection, intergenic reads and usable bases. The resulting cohort comprised 370 participants: 255 with Parkinson’s disease (PD) and 115 healthy controls (HC). Participants were divided into a Training Set (n = 296; 204 PD and 92 HC) and a PPMI Locked Test Set (n = 74; 51 PD and 23 HC). Conventional transcriptomic feature selection and low-expression signature construction were pursued in parallel. The workflow progressed from a 35-gene panel plus the composite low-expression score (LowExpr_score), through a 41-input candidate stage, to a Primary Model containing 28 genes and LowExpr_score. The frozen low-expression reference contained 622 rows, with 322 active contributing genes after pruning; the score remained a single model input. An expanded pool of 95 candidates was evaluated before constrained refinement, yielding the final ORIGIN-PD classifier with 34 gene features and LowExpr_score. Candidate expansion combined extension candidates with mechanism- and literature-informed add-back candidates; it should not be interpreted as exclusively protein–protein-interaction-derived. The PPMI Locked Test Set was not used for coefficient estimation, lambda tuning or threshold fitting, but historical results from this cohort informed model-version selection.

**Table 1.**
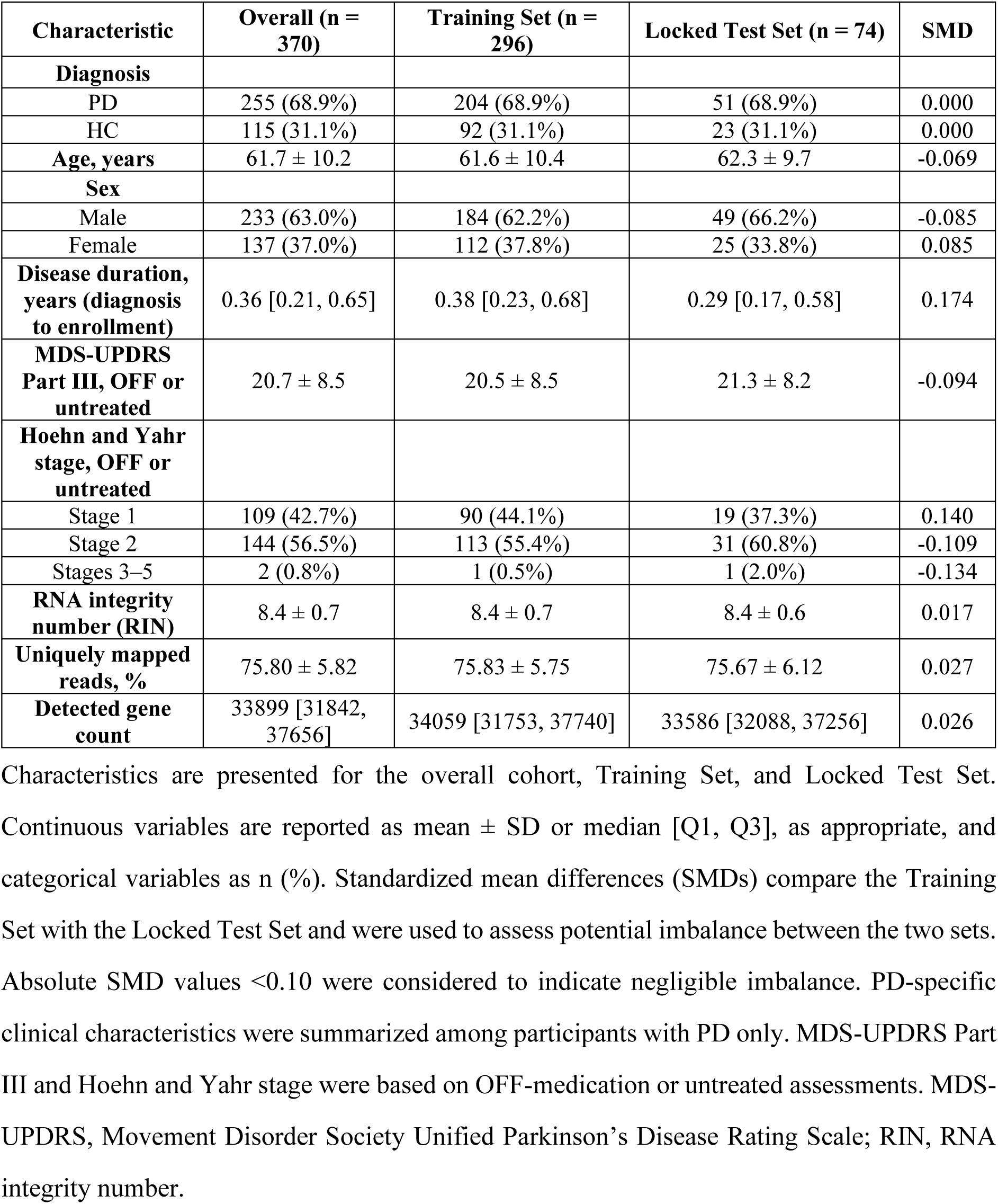
Baseline characteristics of the PPMI cohort. Characteristics are presented for the overall cohort, Training Set, and Locked Test Set. Continuous variables are reported as mean ± SD or median [Q1, Q3], as appropriate, and categorical variables as n (%). Standardized mean differences (SMDs) compare the Training Set with the Locked Test Set and were used to assess potential imbalance between the two sets. Absolute SMD values <0.10 were considered to indicate negligible imbalance. PD-specific clinical characteristics were summarized among participants with PD only. MDS-UPDRS Part III and Hoehn and Yahr stage were based on OFF-medication or untreated assessments. MDS-UPDRS, Movement Disorder Society Unified Parkinson’s Disease Rating Scale; RIN, RNA integrity number.

Transcriptomic feature construction integrated three complementary sources of information. First, 43 genes curated from published literature were assigned to eight functional modules (**Table S1**). Second, DEGs between PD and HC were identified using limma (covariates: diagnosis, sex, sequencing plate, age, and usable sequencing bases) after TMM normalization and log2CPM transformation (prior count = 1), with empirical Bayes moderation (trend and robust). After filtering for expression abundance (CPM ≥1 in at least 10% of all quality-control-passing samples), 21,785 genes remained. Genes with nominal P < 0.05 and absolute log2 fold change > 0.1 were retained (1,044 candidates), ranked by ascending nominal *P* value, descending absolute log2 fold change, and ascending adjusted *P* value. The top 100 unique genes underwent repeated stability assessment, incremental evaluation, and backward elimination, yielding a 35-gene DEG-derived panel. Third, a low-expression feature branch was developed to capture reproducible signals among low-abundance transcripts. In the Training Set, 14,994 low-expression candidates (transcripts with CPM >0.1 in at least 10% of samples but CPM >1 in fewer than 10% of samples) were identified. Diagnosis-stratified subsampling defined a stable 622-gene reference set (228 PD-upregulated and 394 PD-downregulated genes) using the following criteria: *P*<0.05 and **|log2FC|** >0.10 in at least 50% of subsamples, ≥80% directional consistency, and |median log_2_FC| ≥0.10. The 622 genes were further ranked using a prespecified gene-level stability score, and repeated cross-validation within the Training Set supported a pruning depth of 300 without materially reducing PD discrimination, leaving 322 active contributors. Their expression values were combined into a composite variable, termed LowExpr_score, which entered the model as one feature input.

### Performance of the Primary Model

An initial broad screening jointly evaluated the 43 prespecified module-associated genes (**Table S1**) with the full DEG candidate pool (1,044 genes). However, none of the 43 prespecified module-associated genes was consistently retained in this joint multivariable screening, suggesting that their individual signals were insufficiently stable within the broad and heterogeneous candidate space. The transcriptomic features were therefore refined through separate data-driven procedures, yielding a 35-gene DEG-derived panel and the composite LowExpr_score, which together formed a 36-input feature set. On this more compact transcriptomic background, a focused biologically informed evaluation was then performed using nine investigator-specified genes with strong prior relevance to PD and midbrain dopaminergic biology: *NPM1* (our unpublished data)*, WNT1, CTNNB1, LEF1, BDNF, PITX3, NRP2, LMX1B,* and *MSX1*. These genes were evaluated jointly with these existing inputs using repeated fivefold cross-validation in the Training Set. Five genes—*NPM1, WNT1, BDNF, LMX1B*, and *MSX1*—met the predefined selection-frequency criterion of ≥60% across 50 repeated fivefold cross-validation runs and were retained, yielding a 41-input candidate space comprising 40 gene-level features and LowExpr_score for LASSO-based development of the Primary Model (**Figure 1**). Among these five key developmental genes, NPM1 (our unpublished data), LMX1B, and MSX1 are transcriptional regulators of WNT1 expression, whereas BDNF is a downstream target of the WNT1 signaling pathway [8, 28, 32, 52].

Building on the 41-input candidate space established above, LASSO regularization in the Training Set retained 29 non-zero inputs, comprising 28 gene features (*MAP4K4, COL9A3, SRPK1, GSTZ1, CDK6, SOD2, WNT1, NPM1, MSX1, LMX1B, BDNF, NRL, TOR1B, NRM, TMEM45B, OLAH, CACNB3, SNHG11, CD300LD, LINC00623, WDR86-AS1, CD8B2, CEBPA-DT, PTGES3P1, lnc-AMDHD1-1, lnc-FBXO25-2, lnc-TBPL2-6* and *AC007298.1*) and LowExpr_score. The resulting 29-input classifier was designated the Primary Model.

ROC analysis of this Primary Model yielded an apparent Training Set AUC of 0.961 (95% CI, 0.942–0.981) and a PPMI Locked-Test Set AUC of 0.904 (95% CI, 0.832–0.976; **Figure 2A**). The participant-level probability distributions showed higher assigned probabilities in PD than HC, but residual overlap and classification errors remained in both cohorts (**Figure 2B**). At the threshold of 0.69, the Locked Test Set confusion matrix contained 43 true positives, 19 true negatives, four false positives and eight false negatives, corresponding to sensitivity 0.843 and specificity 0.826 (**Figure 2C**). Complementary performance and calibration assessment showed a PR-AUC of 0.954 and a Brier score of 0.115 in Locked Test Set, with a calibration-in-the-large of −0.33 and a calibration slope of 1.13 (**Figure 2D**). The corresponding log-loss was 0.361. In the Training Set, the apparent PR-AUC, Brier score, and log-loss were 0.982, 0.080, and 0.273, respectively (**Figure 2D**). Taken together, these results indicate that the Primary Model provided a strong baseline level of discrimination and calibration in the Locked Test Set, while the residual performance gap relative to the Training Set highlighted scope for further refinement and motivated subsequent model expansion.

**Fig. 2.**
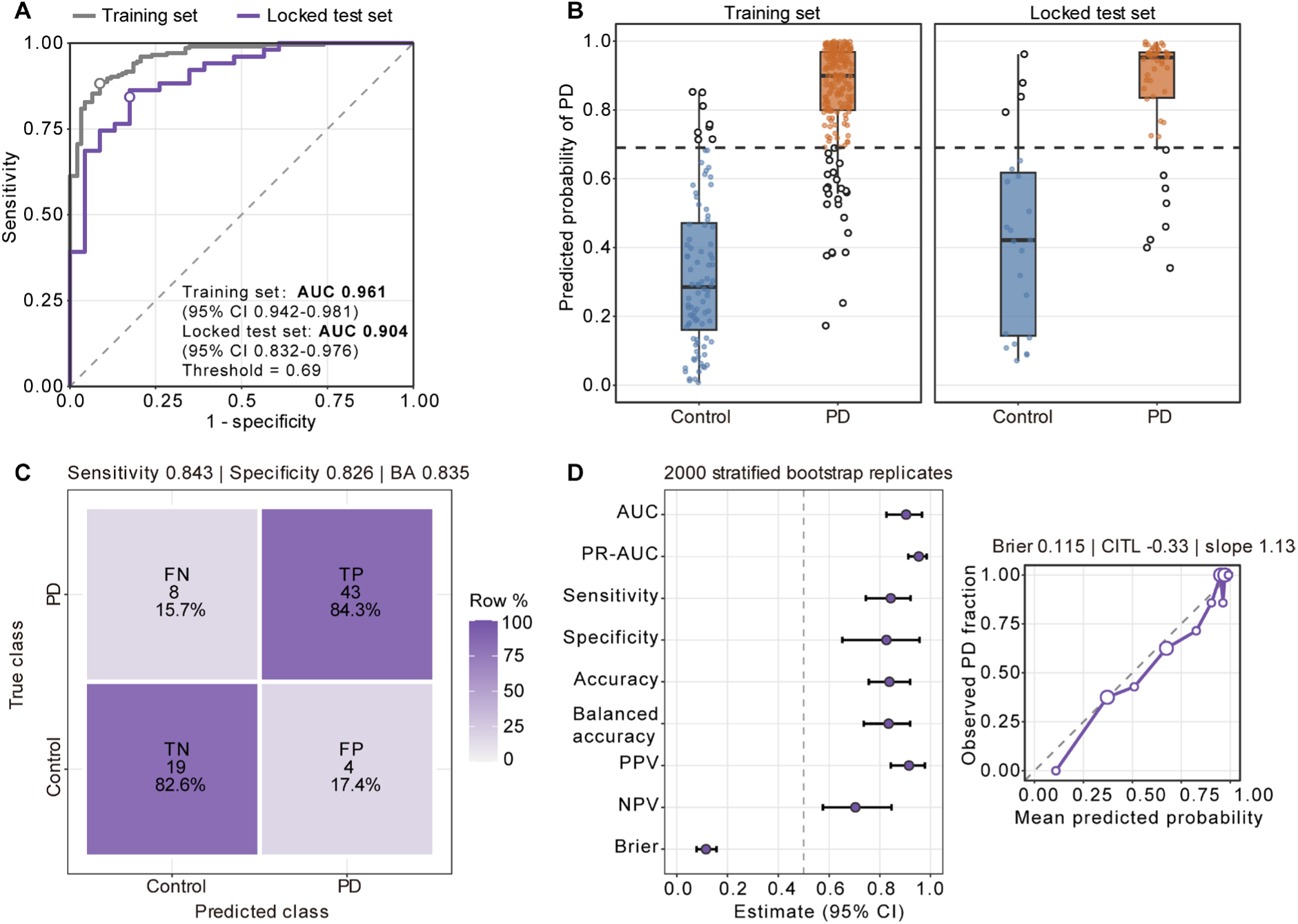
Development and internal evaluation of the Primary Model show high discrimination in the Training Set and PPMI Locked Test Set. The Primary Model comprised 28 gene features and LowExpr_score. (**A**) Receiver operating characteristic (ROC) curves in the Training Set (n = 296; 204 PD and 92 HC) and PPMI Locked Test Set (n = 74; 51 PD and 23 HC). Areas under the ROC curve (AUCs) were 0.961 and 0.904, respectively; annotated 95% confidence intervals (CIs) were calculated using the DeLong method. Open circles mark the operating points at the probability threshold of 0.69. (**B**) Model-assigned PD probabilities by diagnosis and cohort. Points represent participants; open black circles identify classification errors. Boxes indicate the median and interquartile range, and whiskers extend to the most extreme observations within 1.5 times the interquartile range. The dashed horizontal line marks the threshold. (**C**) PPMI Locked Test Set confusion matrix: 43 true positives, 19 true negatives, 4 false positives and 8 false negatives. Percentages are calculated within each true-class row. (**D**) Locked Test Set performance estimates with 95% percentile CIs from 2,000 outcome-stratified bootstrap replicates (left), and quantile-binned calibration (right). Calibration points compare mean assigned probability with observed PD frequency; point size represents the number of participants per bin, and the dashed diagonal denotes ideal calibration. Development performance is apparent performance, not cross-validated performance. Historical Locked Test Set exposure during model-version selection is described in Figure 1 and Methods. PR-AUC, area under the precision–recall curve; BA, balanced accuracy; PPV/NPV, positive/negative predictive value; CITL, calibration-in-the-large.

### Candidate expansion and refinement of the ORIGIN-PD Model achieves high diagnostic performance in the locked test set

Although the Primary Model provided strong initial discrimination, residual overlap between PD and HC and the remaining classification errors indicated scope for further refinement. To broaden the candidate space, model development returned to the 36-input integrated feature set established before LASSO selection. An additional 50 biologically informed candidate genes and previous 9 key developmental genes (**Table S3**) were together introduced on the basis of prior evidence implicating them in PD-relevant biological processes, including mDA neuron identity/development, inflammation and immunity, proteostasis, mitochondrial and oxidative-stress pathways, and microglial or glial responses, together with genes supported by PPI relationships with features in the original 36-input set. Together, these candidates expanded the feature space to 95 inputs. LASSO-based selection within the Training

Set retained 35 non-zero inputs, comprising 34 gene-level features (**Table 2**) and LowExpr_score, which constituted the Intermediate Model. Of the original 35 DEG-derived gene features, 15 were retained: *NRM, TMEM45B, LINC00623, CD8B2, OLAH, TOR1B, CACNB3, AC004908.2, COL9A3, WDR86-AS1, CD300LD, CDK6, GSTZ1, SRPK1,* and *SNHG11*. The remaining 20 original gene features were replaced by 19 features selected from the expanded candidate space: *AGO2, COL9A1, MAPT, WNT1, MSX1, LMX1B, HSPA8, LTA4H, TUBB, CEACAM20, ERRFI1, PRAME, CEACAM18, CEACAM3, HSPD1, NCAM1, SFN, TUBB8,* and *RAP1GAP*. LowExpr_score was retained as the composite low-expression input. Relative to the Primary Model, 18 of the 28 gene features were retained, 10 were removed, and 16 newly selected genes were incorporated (**Figure 3A**).

**Fig. 3.**
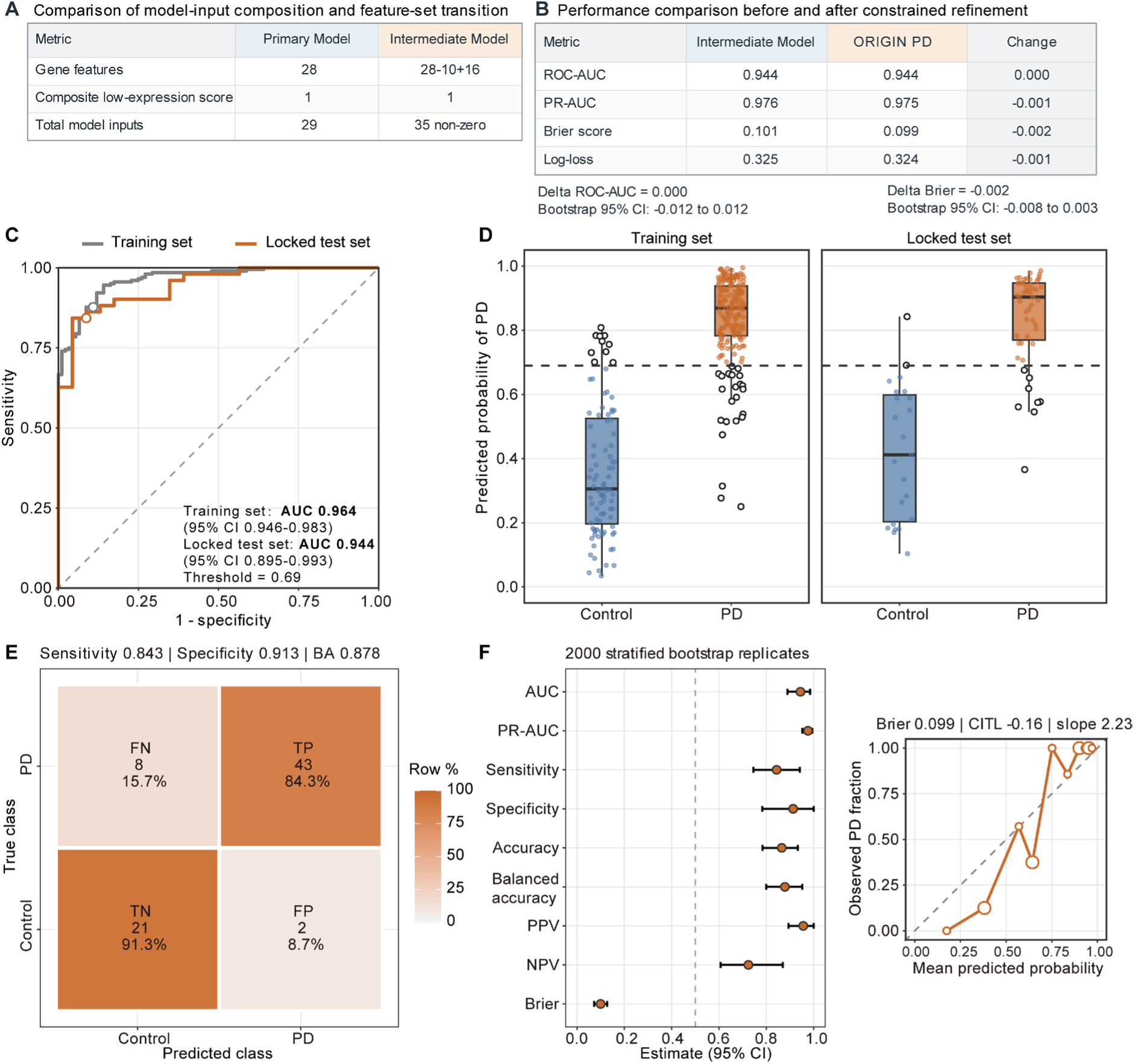
Candidate expansion, constrained refinement and performance of the ORIGIN-PD yield high diagnostic performance in development and locked test sets. (**A**) Input composition before and after candidate expansion. Relative to the Primary Model, 10 of 28 gene features were removed and 16 were added, producing an intermediate classifier with 34 genes and LowExpr_score (total 35 inputs). (**B**) PPMI Locked Test Set performance before and after constrained refinement. *HSPA8* was replaced by *NPM1*, with the other 33 gene coefficients and the LowExpr_score coefficient held fixed; only the *NPM1* coefficient and intercept were re-estimated in the Training Set. Differences are final minus intermediate estimates. The displayed CIs for differences in ROC-AUC and Brier score were obtained from 2,000 paired outcome-stratified bootstrap replicates. The comparisons do not establish a statistically supported improvement in performance. (**C**) Final-classifier ROC curves in the Training Set (n = 296; 204 PD and 92 HC; AUC 0.964) and PPMI Locked Test Set (n = 74; 51 PD and 23 HC; AUC 0.944), with annotated DeLong 95% CIs. Open circles mark the operating points at the frozen threshold of 0.69. (**D**) Assigned PD probabilities by diagnosis and cohort. Boxes show medians and interquartile ranges; whiskers extend to observations within 1.5 times the interquartile range. Points represent participants, open black circles indicate classification errors, and dashed lines mark the threshold. (**E**) Locked Test Set confusion matrix: 43 true positives, 21 true negatives, 2 false positives and 8 false negatives, corresponding to sensitivity 0.843 and specificity 0.913. Percentages are within true-class rows. (**F**) Locked Test Set metric estimates with 95% percentile CIs from 2,000 outcome-stratified bootstrap replicates (left), and quantile-binned calibration (right). Calibration-point size indicates bin size; the diagonal represents ideal calibration. Development estimates are apparent, and Locked Test Set estimates retain the historical model-version-selection limitation described in Methods. PR-AUC, precision–recall AUC; BA, balanced accuracy; PPV/NPV, positive/negative predictive value; CITL, calibration-in-the-large.

**Table 2.** Biological annotation and model characteristics of the 34 gene predictors retained in ORIGIN-PD. The table lists the gene symbol, Ensembl feature ID, gene biotype, and a brief summary of the established or proposed biological role of each gene-level feature included in ORIGIN-PD.

| Gene symbol | Feature ID | Biotype | Biological role |
| --- | --- | --- | --- |
| *LMX1B* | ENSG00000136944 | protein_coding | LIM-homeodomain transcription factor involved in midbrain dopaminergic development. |
| *WNT1* | ENSG00000125084 | protein_coding | Secreted Wnt signalling ligand. |
| *MSX1* | ENSG00000163132 | protein_coding | Homeodomain transcription factor involved in dopaminergic neuronal differentiation. |
| *NPM1* | ENSG00000181163 | protein_coding | Nucleolar phosphoprotein involved in ribosome biogenesis and nucleolar organization. |
| *CEACAM20* | ENSG00000273777 | protein_coding | CEACAM-family protein. |
| *CEACAM3* | ENSG00000170956 | protein_coding | Granulocyte phagocytic receptor mediating Src/Rac-linked uptake of human-adapted bacteria. |
| *CD8B2* | ENSG00000254126 | protein_coding | Gene-specific physiological function remains insufficiently resolved; CD8B function is not assigned by homology. |
| *CD300LD* | ENSG00000204345 | protein_coding | Neutrophil-associated receptor studied in tumour-induced myeloid immunosuppression. |
| *SFN* | ENSG00000175793 | protein_coding | Stratifin/14-3-3 sigma, a signaling scaffold involved in cell-cycle regulation. |
| *TMEM45B* | ENSG00000151715 | protein_coding | Transmembrane protein studied in cancer-cell proliferation and survival. |
| *CEACAM18* | ENSG00000213822 | protein_coding | CEACAM-family protein; PD-specific molecular function remains unestablished. |
| *COL9A1* | ENSG00000112280 | protein_coding | Type IX collagen alpha-1 chain. |
| *COL9A3* | ENSG00000092758 | protein_coding | Type IX collagen alpha-3 chain, an extracellular-matrix component. |
| *GSTZ1* | ENSG00000100577 | protein_coding | Glutathione transferase zeta / maleylacetoacetate isomerase. |
| *OLAH* | ENSG00000152463 | protein_coding | Oleoyl-ACP hydrolase participates in fatty-acid production. |
| *LTA4H* | ENSG00000111144 | protein_coding | Leukotriene-metabolizing enzyme; inhibition has been studied in age-associated memory impairment. |
| *ERRFI1* | ENSG00000116285 | protein_coding | ERBB receptor feedback inhibitor. |
| *HSPD1* | ENSG00000144381 | protein_coding | Mitochondrial HSP60 chaperonin supporting protein folding and mitochondrial proteostasis. |
| *TOR1B* | ENSG00000136816 | protein_coding | TorsinB is an AAA+ protein localized to ER-associated structures. |
| *AGO2* | ENSG00000123908 | protein_coding | Argonaute RNA-silencing effector. |
| *MAPT* | ENSG00000186868 | protein_coding | Microtubule-associated protein tau. |
| *SRPK1* | ENSG00000096063 | protein_coding | Serine/arginine-rich splicing-factor kinase. |
| *NCAM1* | ENSG00000149294 | protein_coding | Neural cell adhesion molecule/CD56, also expressed on natural killer cells. |
| *CDK6* | ENSG00000105810 | protein_coding | Cyclin-dependent kinase involved in cell-cycle control. |
| *TUBB* | ENSG00000196230 | protein_coding | Class I beta-tubulin structural microtubule component. |
| *CACNB3* | ENSG00000167535 | protein_coding | Calcium voltage-gated channel auxiliary beta-3 subunit. |
| *TUBB8* | ENSG00000261456 | protein_coding | Primate-specific beta-tubulin isotype with demonstrated oocyte spindle functions. |
| *RAP1GAP* | ENSG00000076864 | protein_coding | GTPase-activating protein regulating RAP1 signaling. |
| *WDR86-AS1* | ENSG00000243836 | lncRNA | Antisense long non-coding RNA studied in a WDR86-AS1/miR-10b-3p/LITAF regulatory network in preeclampsia models. |
| *LINC00623* | ENSG00000226067 | lncRNA | Long non-coding RNA; a specific PD mechanism has not been established. |
| *AC004908.2* | ENSG00000272812 | lncRNA | Uncharacterized long non-coding transcript; no gene-specific biological function assigned. |
| *SNHG11* | ENSG00000174365 | lncRNA | Long non-coding RNA investigated in hippocampal neurogenesis and plasticity. |
| *NRM* | ENSG00000137404 | protein_coding | Nurim is a multispinning inner nuclear membrane protein. |
| *PRAME* | ENSG00000185686 | protein_coding | Tumour antigen reported to repress retinoic-acid receptor signalling in cancer models. |

Before refinement, ROC analysis of the Intermediate Model yielded apparent Training Set and Locked Test Set AUCs of 0.965 and 0.944, respectively (**Figure S1A**). Its probability distributions retained overlap between diagnoses despite higher assigned probabilities in PD (**Figure S1B**). At the fixed threshold, the Locked Test Set confusion matrix contained 43 true positives, 22 true negatives, one false positive and eight false negatives, with sensitivity 0.843 and specificity 0.957 (**Figure S1C**). The Intermediate Model performance and calibration assessment yielded a Brier score of 0.101, with a calibration-in-the-large of −0.17 and a calibration slope of 2.19 (**Figure S1D**).

Although the Intermediate Model achieved strong discrimination, it was surprising that *NPM1* was excluded from its feature set. NPM1 has been demonstrated by our unpublished data as an important player in the development and survival of SNc DA neurons, and was retained in the Primary Model with a relatively high contribution to model predictions, indicating that it carried informative signal within the original feature space. We therefore investigated this discrepancy and found that *NPM1* was replaced by *HSPA8*, likely because the two proteins physically interact in the nucleus and their expression was strongly correlated in both the Training Set and the Locked Test Set (Spearman’s ρ = 0.845 and 0.799, respectively; **Figure S2**). This interaction is biologically plausible because both NPM1 and HSPA8 are involved in nuclear and nucleolar homeostasis[53]. NPM1 (B23) is a multifunctional nuclear chaperone involved in ribosome biogenesis, chromatin remodeling, and transcriptional regulation, while HSPA8 (Hsc70) is a constitutively expressed molecular chaperone that shuttles between the cytoplasm and the nucleus/nucleoli under stress and plays a central role in protein quality control and chaperone-mediated autophagy[53]. Beyond the nucleolus, HSPA8 also functions in the cytoplasm, lysosomal membrane, and proteasomal degradation pathways[53, 54], and its downregulation by rotenone has been linked to α-synuclein toxicity in PD models[55]. Consistent with a vital role for NPM1 in mDA neurons and PD (our unpublished data), reduced NPM1 mRNA expression has been observed in the SN of PD patients along with disease progression[56]. Since there are evidences supporting NPM1’s critical role in PD progression and its potential as a therapeutic target, we re-evaluated NPM1 as a substitute for HSPA8 in the Intermediate Model to establish a mechanism-informed refinement Model.

A constrained replacement strategy was therefore applied in which *HSPA8* was replaced by *NPM1*, while the coefficients of the remaining 33 gene features and LowExpr_score were held fixed. Only the intercept and *NPM1* coefficient were re-estimated in the Training Set, thereby minimizing perturbation of the established multivariable structure. This refinement yielded the final 35-input ORIGIN-PD, comprising 34 gene-level features (**Table 2**) and LowExpr_score.

Performance was then compared directly before and after constrained refinement in the Locked Test Set (**Figure 3B**). ROC-AUC remained unchanged at 0.9437, with a paired final-minus-intermediate difference of 0.0000 (95% bootstrap CI, −0.0119 to 0.0119). The Brier score decreased slightly from 0.1014 to 0.0992 (difference, −0.00215; 95% CI, −0.00798 to 0.00288), while log-loss decreased from 0.3253 to 0.3242 (difference, −0.00107; 95% CI, −0.01432 to 0.01095). PR-AUC changed minimally from 0.9763 to 0.9754.

ROC evaluation of the ORIGIN-PD yielded an apparent Development AUC of 0.964 (95% CI, 0.946–0.983) and the Locked Test Set AUC of 0.944 (95% CI, 0.895–0.993; **Figure 3C**). ORIGIN-PD assigned higher probabilities to participants with PD than to HC, although some individual observations remained on the incorrect side of the classification threshold (**Figure 3D**). The Locked Test Set confusion matrix contained 43 true positives, 21 true negatives, two false positives and eight false negatives, corresponding to sensitivity 0.843 and specificity 0.913 (**Figure 3E**). The final performance and calibration assessment showed Locked Test Set PR-AUC 0.975, Brier score 0.0992, accuracy 0.865 and balanced accuracy 0.878, with calibration-in-the-large −0.16 and calibration slope 2.23 (**Figure 3F**). Locked Test Set log-loss in the saved results was 0.324. Corresponding apparent Development PR-AUC, Brier score and log-loss were 0.984, 0.0835 and 0.294; sensitivity and specificity were 0.877 and 0.891.

Relative to the Primary Model, ORIGIN-PD showed a numerically higher AUC in the Locked Test Set (0.944 vs 0.904) and improved specificity (0.913 vs 0.826), while sensitivity remained unchanged and the number of false positives decreased from four to two. In contrast, comparison with the Intermediate Model showed essentially unchanged discrimination, although false positives increased slightly from one to two. Thus, the transition to ORIGIN-PD primarily reflected a refinement of feature composition rather than a uniform improvement across all performance metrics. Importantly, the constrained refinement preserved the strong predictive performance of the Intermediate Model while incorporating *NPM1* on the basis of convergent model-derived, biological, and expression-correlation evidence. Overall, these findings support ORIGIN-PD as a biologically informed refinement that maintained robust discrimination while improving the mechanistic coherence of the final feature set, rather than as a model with statistically superior performance across all metrics.

### Biological interpretation of ORIGIN-PD features and module contributions reveals etiologic heterogeneity with potential for PD stratification

Several retained features have established or emerging links to PD biology. *MAPT* is a well-established PD susceptibility locus[57], while *CDK6* and *AGO2* have been implicated in PD-related neuronal dysfunction and α-synuclein biology, respectively[58, 59]. The mDA neuron identity-related genes *WNT1, MSX1, NPM1* and *LMX1B* are key regulators of mDA neuron development and maintenance[60–62] (our unpublished data), whereas *NPM1* has been reported to be altered in the SN DA neurons of patients with PD[56], consistent with our recent findings (our unpublished data) implicating the NPM1/WNT1 axis in mDA neuron development and survival. Several additional features (*NRM, RAP1GAP, COL9A3, LINC00623,* and *TOR1B*) identified by ORIGIN-PD have also been reported in previous blood-transcriptomic studies of PD, providing convergent support for the reproducibility and biological plausibility of these signals[63].

To determine whether the predictive structure of ORIGIN-PD could be related to biologically interpretable signals, we examined the retained features at both the individual-gene and functional-module levels, integrating model coefficients, marginal PD–HC expression differences, SHAP-based contributions, and predefined biological annotations. This framework was used to distinguish the conditional contribution of each feature within the multivariable model from its marginal expression pattern between diagnostic groups. Coefficient-level inspection identified both PD-directed and HC-directed conditional associations among the retained inputs; *NPM1* had a positive frozen coefficient of +0.747 (**Figure 4A**). The pooled feature-distribution analysis showed that conditional directions were not interchangeable with marginal expression differences: *NPM1* had a negative pooled Hedges’ g of −0.179 across the 370 participants despite its positive coefficient (**Figure 4B**).

**Fig. 4.**
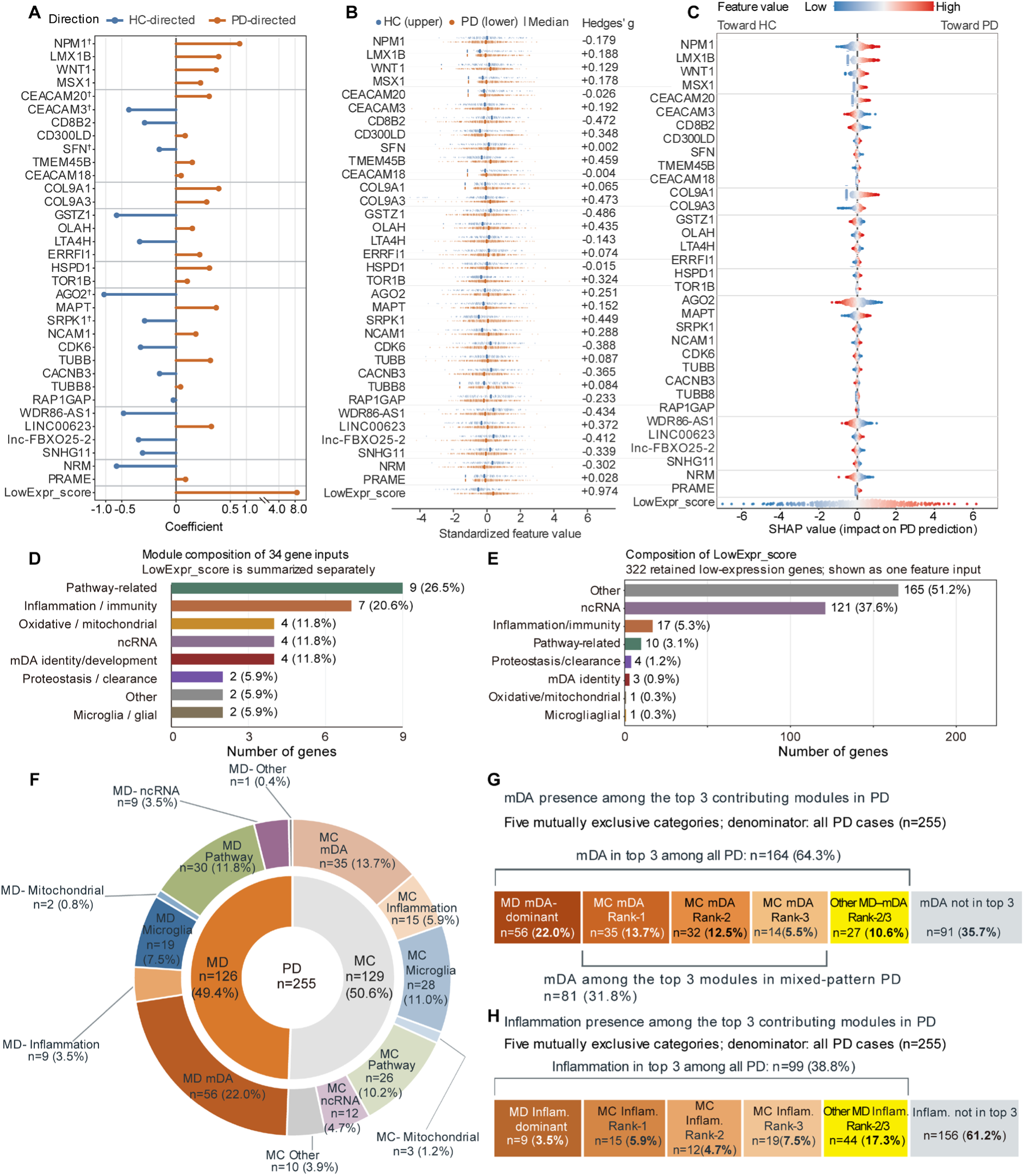
Conditional model contributions and marginal PD-HC feature differences reveal module-level biological heterogeneity in the ORIGIN-PD. (**A**) Frozen coefficients for the 34 gene inputs and LowExpr_score on their original model-input scales. Orange and blue indicate positive and negative coefficients, respectively; the broken axis accommodates the LowExpr_score coefficient. These coefficients describe conditional predictive associations, not marginal expression differences. Daggers (^†^) identify Training Set marginal–conditional direction discordance. (**B**) Feature distributions pooled across the Training Set and PPMI internal Locked Test Set (n = 370; 255 PD and 115 HC), standardized using the Development reference mean and standard deviation. Dots represent participants, with HC above and PD below each feature row; vertical ticks indicate group medians. The adjacent Hedges’ g values describe pooled marginal PD–HC differences, with positive values indicating higher values in PD. (**C**) Analytical SHapley Additive exPlanations (SHAP) values on the logit scale. Positive and negative values shift the model output toward PD and HC, respectively. Colour represents Development-standardized feature values; the colour scale is clipped at −2 and +2, without clipping SHAP values. Saved values for both cohorts are pooled only for visualization, retaining the same Development background. Features are grouped by module and ordered within modules using Development mean absolute SHAP; LowExpr_score is shown separately. (**D**) Module composition of the 34 gene inputs: pathway-related (9), inflammation/immunity (7), oxidative/mitochondrial (4), non-coding RNA (4), midbrain dopaminergic (mDA) neuron identity (4), proteostasis/clearance (2), microglia/glial (2) and other (2). (**E**) Module composition of the 322 active low-expression genes. These genes collectively contribute to one composite input and are distinct from the 622-row reference specification. (**F**) Distribution of functional-module contribution patterns among all 255 participants with PD. The inner ring distinguishes monomodule-dominant patterns (MD; n = 126) from multimodule-contribution (MC; n = 129), and the outer ring indicates the first-ranked functional module for each participant. (**G**) mDA contribution patterns among all 255 PD participants. mDA was the dominant module in 56 MD cases. Among the 129 MC cases, mDA ranked first in 35, second in 32, and third in 14 participants. A further 27 MD participants had mDA ranked second or third, whereas mDA was outside the top three in 91 participants. Overall, mDA ranked among the top three modules in 164/255 participants 64.3%), including 81/129 MC cases (62.8%). (**H**) Corresponding inflammation/immunity contribution patterns. Inflammation/immunity was dominant in 9 MD cases and ranked first, second, or third in 15, 12, and 19 MC cases, respectively. A further 44 MD participants had inflammation/immunity ranked second or third, whereas it was outside the top three in 156 participants. Overall, inflammation/immunity ranked among the top three modules in 99/255 participants (38.8%).

The participant-level SHAP analysis identified LowExpr_score as the input with the largest mean absolute contribution in both cohorts (**Figure 4C**). Among gene features, *AGO2, COL9A1, LMX1B* and *NPM1* had the largest Development mean absolute SHAP values. The accompanying saved importance comparison showed concordant rankings across the 35 inputs (Spearman ρ = 0.980), with all ten Training Set top-ranked features also in the Locked Test Set top ten. The underlying cohort-specific direction audit further distinguished marginal and conditional effects. Among the 34 gene features, 28 showed concordant Training marginal, univariate and frozen multivariable directions, whereas six (*SRPK1, AGO2, CEACAM20, SFN, CEACAM3* and *NPM1*) showed marginal–conditional sign reversals. Marginal direction replicated from Training to Lock Test for 26 genes, and cohort-specific Hedges’ g estimates were correlated (Spearman ρ = 0.801). *NPM1* expression was lower in PD in both Training Set (g = −0.151) and Lock Test Set (g = −0.281). Overall, the broad concordance between model-derived directions and observed expression differences suggests that ORIGIN-PD largely captures biologically interpretable disease-related signals. For the small subset of genes showing sign reversals, the discrepancy likely reflects the distinction between marginal expression differences and conditional multivariable effects: whereas Hedges’ g summarizes the unadjusted PD–HC difference for an individual gene, the frozen model coefficient represents its contribution after accounting for the other correlated features in the model.

The 34 gene-level inputs were classified according to a prespecified eight-category functional framework: pathway-related (*CACNB3, CDK6, SRPK1, MAPT, AGO2, TUBB, TUBB8, RAP1GAP,* and *NCAM1*; n = 9), inflammation/immunity (*TMEM45B, CEACAM3, SFN, CD300LD, CEACAM18, CD8B2*, and *CEACAM20*; n = 7), oxidative/mitochondrial (*GSTZ1, LTA4H, ERRFI1*, and *OLAH*; n = 4), non-coding RNA (*SNHG11, LINC00623, WDR86-AS1*, and *AC004908.2*; n = 4), proteostasis/clearance (*TOR1B* and *HSPD1*; n = 2), mDA neuron identity/development (*WNT1, MSX1*, *NPM1* and *LMX1B*; n = 4), microglia/glial (*COL9A3* and *COL9A1*; n = 2), and other (*NRM* and *PRAME*; n = 2; **Figure 4D**). The composition analysis of the 322 active low-expression genes showed a different distribution, dominated by other/unassigned genes (165; 51.2%) and non-coding RNAs (121; 37.6%; **Figure 4E**). The remaining categories comprised inflammation/immunity (17), pathway-related (10), proteostasis/clearance (4), mDA neuron identity/development (3), oxidative/mitochondrial (1) and microglia/glial (1). The participant-level module classification separated the 255 PD participants into 126 (49.4%) mono-module-dominant (MD) contribution pattern (defined as **≥**30% of the total absolute module contribution) and 129 (50.6%) with multi-module contribution (MC) patterns (**Figure 4F**). Within MD subtype, mDA neuron identity/development was the dominant module in 56 MD participants. Among MC subtype, mDA neuron identity/development module ranked first, second, and third in 35, 32, and 14 cases, respectively. An additional 27 MD participants had mDA neuron identity/development module ranked second or third. Overall, mDA neuron identity/development was among the three highest-ranked modules in 164 of 255 participants (64.3%), including 81 of 129 MC participants (62.8%; **Figure 4G**). For inflammation/immunity, there were 9 MD cases with inflammation/immunity as the dominant module; among MC participants, inflammation/immunity ranked first, second, and third in 15, 12, and 19 cases, respectively. An additional 44 MD participants had inflammation/immunity ranked second or third. Overall, inflammation/immunity was among the top three modules in 99 of 255 participants (38.8%), including 46 of 129 MC participants (35.7%; **Figure 4H**). The most frequent first-ranked contribution patterns were mDA neuron identity/development (35.7%), pathway-related signaling (22.0%), microglial/glial responses (18.4%), and inflammation/immunity (9.4%). These same biological domains were also frequently represented among the top three module contributions across the cohort (**Table 3**). In ORIGIN-PD, proteostasis/clearance-related contributions were broadly retained across PD participants, although they were predominantly positioned at ranks 4-8 rather than within the top three contributors (**Table S4**). In ORIGIN-PD, *HSPA8* was replaced by *NPM1*, with *NPM1* reassigned to the mDA neuron identity/development module. To further examine the impact of this substitution, we compared participant-level module contribution patterns with those of the Intermediate Model, in which *HSPA8* was retained within the proteostasis/clearance module. In the Intermediate Model, the proteostasis/clearance module showed substantially greater representation among the leading individual-level contributors (**Figure S3**). This comparison indicates that the reduced top-three representation of proteostasis/clearance in ORIGIN-PD is largely attributable to the model refinement and feature reassignment rather than to a complete absence of proteostasis-related signal. These findings suggest that ORIGIN-PD may capture biologically distinct contribution patterns across individuals, highlighting its potential to identify PD subgroups with different predominant etiologic or pathogenic signatures.

**Table 3.** Distribution of functional-module contribution ranks among participants with PD. The table summarizes the rank distribution of the eight prespecified functional modules among 255 participants with PD. Modules were ranked for each participant according to the absolute magnitude of their signed module contributions. MD participants (n = 126) are summarized by their dominant rank-1 (R1) module, whereas R1 to R3 distributions are shown separately for MC participants (n = 129) and for the full PD cohort (n = 255). Percentages for strong R1, mixed top-three, and all-PD top-three frequencies were calculated using denominators of 126, 129, and 255, respectively. LowExpr_score and the model intercept were excluded from module ranking.

| Module | MD R1<br>n (% of<br>126) | MC<br>R1 | MC<br>R2 | MC<br>R3 | MC top3<br>n (% of<br>129) | All<br>PD<br>R1 | All<br>PD<br>R2 | All<br>PD<br>R3 | All PD top3<br>n (% of<br>255) |
| --- | --- | --- | --- | --- | --- | --- | --- | --- | --- |
| mDA neuron identity | 56 (44.4%) | 35 | 32 | 14 | 81 (62.8%) | 91 | 52 | 21 | 164 (64.3%) |
| Inflammation<br>/immunity | 9 (7.1%) | 15 | 12 | 19 | 46 (35.7%) | 24 | 30 | 45 | 99 (38.8%) |
| Microglia/glia | 19 (15.1%) | 28 | 32 | 24 | 84 (65.1%) | 47 | 64 | 42 | 153 (60.0%) |
| Proteostasis<br>/clearance | 0 (0.0%) | 0 | 0 | 0 | 0 (0.0%) | 0 | 0 | 0 | 0 (0.0%) |
| Pathway-related | 30 (23.8%) | 26 | 20 | 25 | 71 (55.0%) | 56 | 45 | 51 | 152 (59.6%) |
| Non-coding RNA | 9 (7.1%) | 12 | 15 | 21 | 48 (37.2%) | 21 | 32 | 44 | 97 (38.0%) |
| Other | 1 (0.8%) | 10 | 13 | 14 | 37 (28.7%) | 11 | 22 | 24 | 57 (22.4%) |
| Oxidative<br>/mitochondrial | 2 (1.6%) | 3 | 5 | 12 | 20 (15.5%) | 5 | 10 | 28 | 43 (16.9%) |

Representative profiles were selected to illustrate how different combinations of functional-module contributions could underlie ORIGIN-PD predictions at the individual level (**Figure 5A–H**). The eight examples represented each of the prespecified modules as the first-ranked contributor and showed substantial heterogeneity in the relative magnitude and direction of contributions from the remaining modules, despite consistently high assigned PD probabilities (0.74–0.97). Most selected cases met the predefined MD criteria. Because no oxidative/mitochondrial MD case was available, an MC case in which the oxidative/mitochondrial module was the first-ranked positive contributor was selected as a fallback example. These profiles demonstrate that similar model-assigned PD probabilities can arise from distinct combinations of biologically annotated feature contributions, highlighting inter-individual heterogeneity in the predictive structure of ORIGIN-PD.

**Fig. 5.**
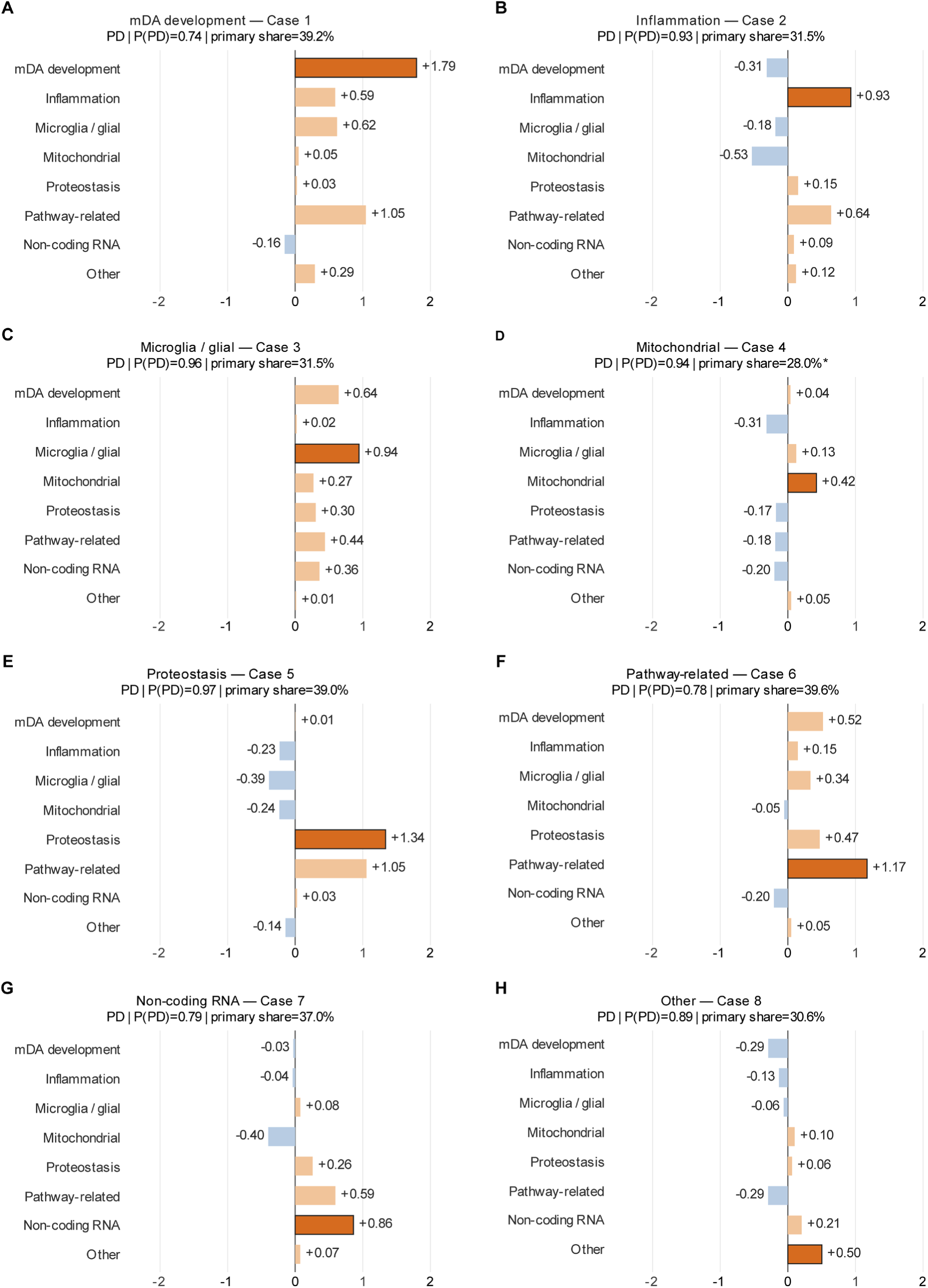
Representative individual module contribution profiles support the potential for etiology-informed stratification of PD. Eight illustrative PD participants from the Training Set are shown, one for each first-ranked module: (**A**) mDA neuron identity/development, (**B**) inflammation/immunity, (**C**) microglia/glial, (**D**) oxidative/mitochondrial, (**E**) proteostasis/clearance, (**F**) pathway-related, (**G**) non-coding RNA and (**H**) other. Bars represent signed module contributions on the logit scale, with positive and negative values indicating contributions toward PD and HC classification, respectively.

### External PD/HC evaluation and non-PD cohort testing confirm robust performance and specificity of ORIGIN-PD

To examine whether the predictive signal captured by ORIGIN-PD could be transferred beyond the PPMI cohort, we next evaluated the frozen classifier in external datasets without model refitting or threshold optimization. These analyses were designed to assess both cross-cohort PD/HC discrimination and the extent to which the classifier generated PD-like predictions in non-PD neurological or systemic disease cohorts.

External evaluation in GSE165082, comprising 12 PD and 14 HC samples, yielded an AUC of 0.667 (95% CI, 0.453–0.881; **Figure 6A**). At the frozen probability threshold of 0.69, four PD and two HC samples were classified as PD-like, corresponding to a sensitivity of 0.333 and a specificity of 0.857 (**Figure 6B**). PR-AUC, Brier score, and log-loss were 0.663, 0.272, and 0.761, respectively. These results indicate that ORIGIN-PD retained some discriminatory signal and relatively high specificity in this external cohort, but sensitivity and overall predictive performance were substantially attenuated compared with the PPMI Locked Test Set. The wide confidence interval and limited number of correctly identified PD cases also indicate considerable uncertainty. This attenuation may reflect the small sample size, differences in cohort composition, assay or preprocessing characteristics, and cross-dataset expression-distribution shifts. In addition, several sample-level sequencing-quality metrics used for PPMI QC were not available in the public GSE165082 dataset, preventing full reproduction of the PPMI QC procedure and limiting assessment of sample comparability. Performance may also have been affected by application of the frozen model and threshold without dataset-specific recalibration. Accordingly, GSE165082 provides evidence of partial cross-cohort transferability but also highlights the need for validation in larger, fully independent external PD cohorts.

**Fig. 6.**
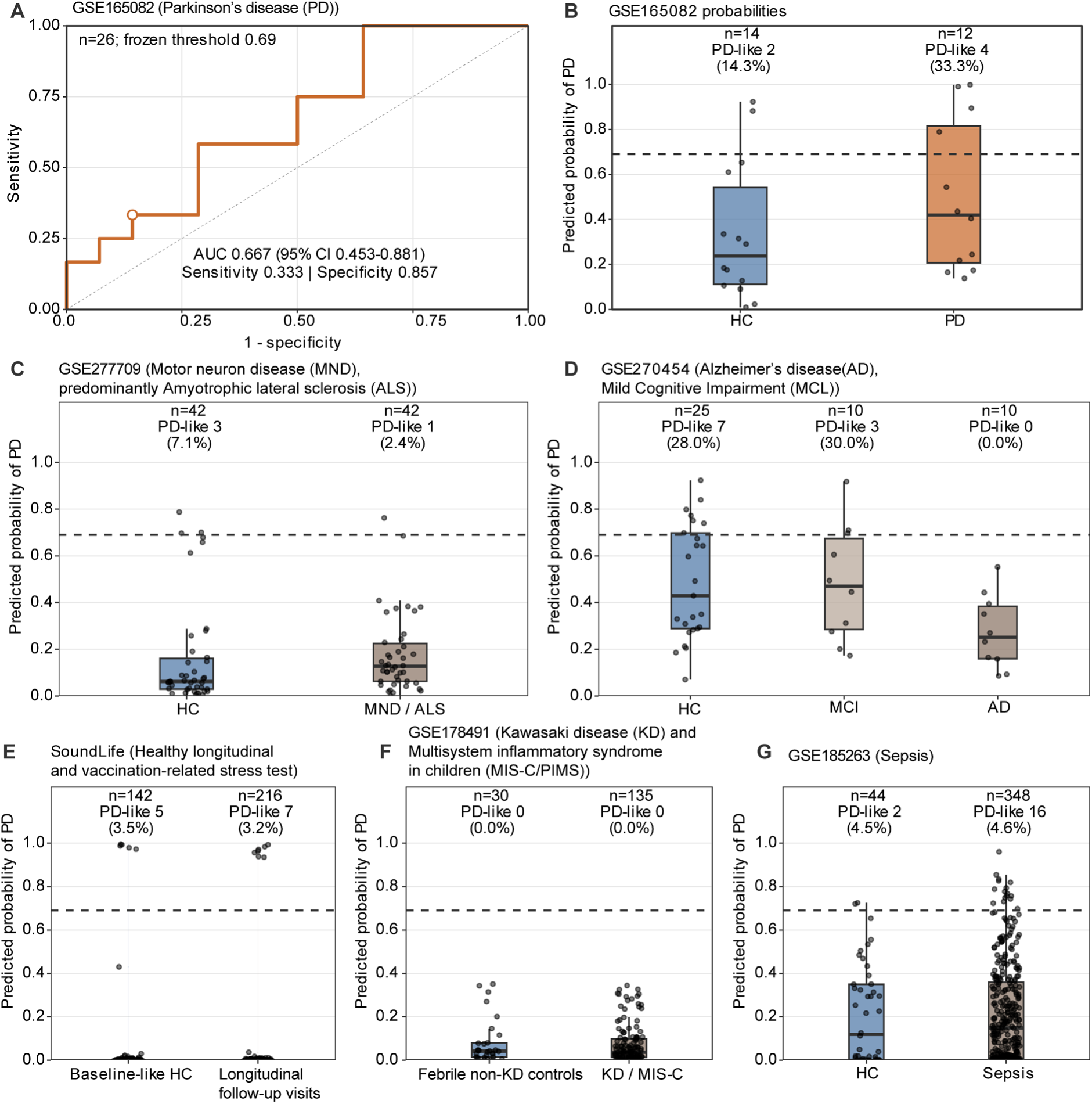
External projections and disease-control stress tests of ORIGIN-PD. The final ORIGIN-PD was evaluated using its frozen coefficients and a probability threshold of 0.69, without external-cohort refitting or threshold optimization. (**A**) ROC curve for GSE165082 (12 PD and 14 HC), with AUC 0.667 and a DeLong 95% CI of 0.453–0.881. The open circle indicates sensitivity 0.333 and specificity 0.857 at the frozen threshold. (**B**) GSE165082 assigned probabilities; 4 of 12 PD and 2 of 14 HC observations exceeded the threshold. (**C**) GSE277709 neurological disease-control stress test, comprising 42 HC and 42 motor-neuron-disease/amyotrophic-lateral-sclerosis observations; 3 and 1, respectively, exceeded the threshold. (**D**) Exploratory GSE270454 sensitivity analysis comprising 25 HC, 10 mild cognitive impairment (MCI) and 10 Alzheimer’s disease observations; 7, 3 and 0, respectively, exceeded the threshold. Only 31 of 34 gene inputs and 113 of 322 active low-expression genes were available, so this panel is not a complete-input ORIGIN-PD validation. (**E**) SoundLife healthy-cohort transferability assessment. Five of 142 selected baseline-like observations and 7 of 216 other observations exceeded the threshold. (**F**) GSE178491 inflammatory disease stress test, comprising 30 comparator observations and 135 observations in the group labelled KD/MIS-C in the figure; none exceeded the threshold. (**G**) GSE185263 sepsis stress test, comprising 44 HC and 348 sepsis observations; 2 and 16, respectively, exceeded the threshold. In B-G, points represent observations; boxes show medians and interquartile ranges, whiskers extend to observations within 1.5 times the interquartile range, and dashed lines mark the threshold. KD, Kawasaki disease; MIS-C, multisystem inflammatory syndrome in children.

The neurological disease-control assessment in GSE277709, comprising 42 patients with sporadic motor neuron disease (MND; 86% classic amyotrophic lateral sclerosis (ALS)) and 42 age-and sex-matched HCs, showed PD-like outputs in three of 42 HC samples (7.1%) and one of 42 motor-neuron-disease samples (2.4%; **Figure 6C**). The low rate of PD-like classification in the motor-neuron-disease group supports good cross-disease specificity, with limited misclassification of non-PD neurodegenerative disease samples as PD.

The neurological disease-control assessment in GSE270454, comprising 25 cognitively asymptomatic controls, ten participants with mild cognitive impairment (MCI), and ten patients with Alzheimer’s disease (AD) dementia, showed threshold-positive outputs in seven of 25 HC samples (28.0%), three of ten mild cognitive impairment samples (30.0%), and none of ten Alzheimer’s disease samples (**Figure 6D**). However, only 31 of 34 ORIGIN-PD gene-level inputs and 113 of 322 active low-expression components were available in this dataset. The relatively high proportion of threshold-positive outputs in the HC and MCI groups suggests reduced stability under substantial feature loss. Notably, none of the Alzheimer’s disease samples were classified as PD-like, supporting good cross-disease specificity against another major neurodegenerative disorder and indicating limited tendency of ORIGIN-PD to misclassify Alzheimer’s disease as PD.

The healthy-cohort transferability assessment in SoundLife included 358 observations from 48 individuals. PD-like outputs were observed in only five of 142 selected baseline-like observations (3.5%) and seven of 216 other observations (3.2%; **Figure 6E**). Although these data included repeated observations rather than 358 independent participants, the consistently low threshold-positive rates across both subsets support stable classifier behavior in a healthy population and suggest a low tendency toward nonspecific PD-like classification.

The inflammatory disease stress test in GSE178491, comprising pediatric patients with Kawasaki disease (KD) and multisystem inflammatory syndrome in children (MIS-C), together with febrile non-KD controls, yielded no threshold-positive outputs among either the 30 comparator observations or the 135 KD/MIS-C observations (**Figure 6F**), indicating that systemic inflammatory states in this cohort did not readily trigger PD-like predictions. Similarly, in GSE185263, which comprised 348 adults presenting with suspected or established sepsis across emergency-room and intensive-care settings and 44 healthy controls, PD-like outputs were observed in only two of 44 HC samples (4.5%) and 16 of 348 sepsis/critical-illness samples (4.6%; **Figure 6G**). Together, these non-PD cohort assessments showed consistently low rates of PD-like classification across healthy, inflammatory, and critically ill populations, supporting good cross-cohort specificity of ORIGIN-PD and limited nonspecific activation outside the PD setting.

## Discussion

Sporadic PD represents a heterogeneous syndrome rather than a single nosological entity, yet no validated framework currently exists to stratify patients according to their predominant pathogenic drivers. Increasing genetic, molecular and biomarker evidence supports substantial biological heterogeneity within clinically defined PD, prompting a shift toward biologically informed classification and precision-medicine frameworks[2, 64–69]. This unmet need has become a major bottleneck for disease-modifying trials, particularly for mechanism-based interventions such as NLRP3 inflammasome or NF-κB inhibitors, where response is likely confined to inflammation-driven subgroups. Accordingly, biomarker-guided screening, stratification and enrichment are increasingly recognized as important components of disease-modifying trial design in PD[70–72]. Here, we present ORIGIN-PD, an interpretable, gene-module-based classifier that assigns each sporadic PD patient a quantitative contribution profile across eight pathobiology-related functional modules.

In a stringently QC-controlled PPMI whole-blood RNA-seq cohort, ORIGIN-PD achieved robust discriminative performance (AUC 0.944, sensitivity 0.843, specificity 0.913). Previous blood transcriptomic classifiers developed using the same PPMI cohort showed more modest and less balanced performance, including an AUC of 0.713 with a sensitivity of 0.817 and specificity of 0.455 in a study by Pantaleo et al. (2022), an AUC of 0.743 with a sensitivity of 0.571 and specificity of 0.824 in a study by Kelly et al. (2023), and an AUC of 0.745 with a sensitivity of 0.318 and specificity of 0.880 in a study by Zhao et al. (2025)[63, 73, 74]. External transferability was not evaluated by Pantaleo et al. and was not assessed beyond an internal train-test split by Kelly et al.; although Zhao et al. performed cross-dataset evaluation, the external cohort had contributed to upstream feature discovery. In comparison, ORIGIN-PD achieved an AUC of 0.944, with a sensitivity of 0.843 and specificity of 0.913 in the Locked Test Set, indicating stronger and more balanced discriminative performance. Notably, our model maintained high specificity against Alzheimer’s disease, amyotrophic lateral sclerosis, and systemic inflammatory conditions, supporting its disease-specificity for PD rather than a generic neurodegeneration or inflammation signature.

Beyond its discriminative performance, many features retained by ORIGIN-PD are supported by prior human PD studies or experimental evidence related to PD biology. Among the 34 retained genes, MAPT has direct human genetic support for PD susceptibility[75], whereas WNT1 has experimentally supported roles in dopaminergic neuronal survival and resistance to toxin-induced injury[60], and LMX1B contributes to autophagic homeostasis and stress resilience in human midbrain dopaminergic neurons[76]. MSX1 additionally contributes to embryonic mDA specification[62]. Other retained features converge on neurodegeneration-related cellular processes: AGO2 has been implicated in ɑ-synuclein biogenesis[58], NPM1 expression and nucleolar localization are reduced in residual SN DA neurons in patients with PD [56] (our unpublished data), and HSPD1 has been associated with dopaminergic injury in human tissue and toxin-based models[77]. CDK6 activity has also been reported to be increased in peripheral cells from patients with PD, while inhibition of the cyclin D3/CDK6 pathway reduced 6-OHDA-induced dopaminergic neuronal injury[59]. Additional retained features have also emerged from human PD molecular studies. *CACNB3* was reported to be downregulated in laser-captured SNc DA neurons from patients with PD, providing direct transcriptomic evidence linking altered calcium-channel regulation to vulnerable DA neurons[78]. More recently, *TUBB* was identified among recurrently dysregulated transcripts in a meta-analysis of PD gene-expression datasets[79], while exonic copy-number loss involving *TUBB8* was reported in a paired analysis of siblings discordant for familial PD, suggesting a potential but as yet unconfirmed genetic association[80]. *ERRFI1* has also been detected as an altered peripheral immune-related transcript in a mouse PD model, although its specific role in PD remains unresolved[81]. The inflammation/immunity features show a broader range of supporting evidence. CEACAM3 promotes granulocyte Syk-CARD9-NF-κB signaling and chemokine release following pathogen engagement[82], whereas phosphorylated CEACAM20 activates Syk-dependent NF-κB signaling and IL-8 production[83]. Stratifin, encoded by *SFN*, also modulates NF-κB signaling by facilitating p65 nuclear export, thereby limiting selected NF-κB responses[84]. In contrast, CD300LD is more directly linked to STAT3-S100A8/A9-mediated myeloid immune regulation[85], while TMEM45B has been associated with inflammation-induced hypersensitivity[86]. *CD8B2* has been reported as differentially expressed in human PD substantia nigra[87], whereas CEACAM18 was identified in an exploratory plasma extracellular-vesicle proteomic analysis of PD[88], providing additional PD-related evidence. Notably, 10 of the 34 gene-level features—*NRM, LINC00623, TMEM45B, TOR1B, COL9A3, CDK6, GSTZ1, LTA4H, RAP1GAP,* and *NCAM1*— were also prioritized in a previous PPMI whole-blood transcriptomic study[63], providing additional support for the robustness and reproducibility of these transcriptomic signals across different analytical approaches. Evidence for several other features is more preliminary. For example, *SNHG11* has been included in exploratory plasma lncRNA profiling in PD, but a gene-specific association with PD has not yet been established[89]. However, features such as *OLAH, AC004908.2, WDR86-AS1, SRPK1, COL9A1*, and *PRAME* remain poorly characterized in PD, suggesting that they may represent underexplored molecular signals and provide new directions for investigating previously unrecognized aspects of PD biology.

The most striking biological finding of this study is the predominance of the mDA neuron identity/development module, comprising the transcription factors *MSX1*, *NPM1*, *LMX1B* and the signaling molecule *WNT1*. These genes belong to a well-established developmental network governing specification and differentiation of mDA neurons. WNT1-dependent signaling establishes and maintains mDA progenitor identity, whereas transcription factors MSX, NPM1 and LMX, participate in dopaminergic fate specification and neurogenesis [27, 28, 62, 90] (our unpublished data). Importantly, these developmental regulators retain functions beyond embryogenesis: LMX1B-dependent programs are involved in autophagic-lysosomal integrity, and sustained LMX1A/B activity is required for mitochondrial function and long-term survival of adult mDA neurons[91, 92]. This module emerged as the single dominant driver (contribution >30%) in 22.0% of patients and ranked among the top three contributors in 62.8% of multi-module contribution cases. Collectively, the mDA neuron identity/development-related gene regulatory pathway was among the top three contributors in 64.3% of the entire sporadic PD cohort.

This observation resonates with our recent mechanistic work demonstrating that the NPM1/WNT1 axis critically governs the number and survival of SNc DA neurons in mice (our unpublished data). Independent neuropathological evidence also supports perturbation of NPM1-related nucleolar biology in PD: reduced NPM1 expression and diminished nucleolar NPM1 immunoreactivity have been reported in the SN of PD patients[56], while recent spatial transcriptomic analyses further revealed downregulation of ribosomal protein synthesis and ubiquitin-proteasome-related pathways in the SN of PD patients, with NPM1 emerging as a key downregulated network node[93]. The convergence of transcriptomic subtyping with independent experimental evidence strongly argues that early-life or developmental-stage deficits in mDA development and maintenance may predispose a substantial proportion of sporadic PD cases, a concept that shifts the traditional emphasis from purely neurodegenerative to neurodevelopmental vulnerability. The broad involvement of the mDA-identity/development module in sporadic PD cases reinforces the possibility of developing NPM1/WNT1-targeted drug for sporadic PD.

Intriguingly, the LASSO-derived model identified *HSPA8* as a predictive feature that carried highly overlapping predictive information with *NPM1*. Far from a spurious computational artifact, this finding has plausible mechanistic underpinnings: HSPA8 (Hsc70) is a constitutively expressed molecular chaperone that physically interacts with NPM1 in the nucleolus, facilitating proper ribosomal RNA processing and coordinating the transcriptional activity of mDA-specification genes[56, 93, 94]. The ability of *HSPA8* expression to recapitulate the predictive information of *NPM1* suggests that the chaperone network supporting nucleolar integrity and WNT signaling is a critical, previously underappreciated determinant of PD susceptibility. Our findings also suggest that HSPA8 is a potential target of PD-modifying therapy too.

Conversely, several well-established PD-or mDA-related genes, including *NR4A2* (NURR1) and *EN1*[95–97], were deliberately included in the prespecified eight-module candidate framework but were not retained in ORIGIN-PD. Their exclusion likely reflects weak or variable peripheral expression and redundancy with correlated transcriptomic signals. Because LASSO retains features according to their independent incremental contribution in a multivariable context, mechanistically important genes such as *NR4A2* or *EN1* may not be selected when they add little unique information in whole-blood data. This also exemplifies the value of transparent, feature-attributable AI — unlike “black-box” deep learning models, ORIGIN-PD’s module-based architecture allows us to trace predictive signals back to biologically coherent pathways, enhancing both interpretability and translational trust.

A second potential implication concerns inflammation-targeted therapies. The inflammation/immunity module includes several genes with links to inflammatory signaling, including NF-κB-related pathways, although the underlying mechanisms are heterogeneous. Moreover, the NLRP3 inflammasome, a core component of innate immunity, has also been implicated in PD progression[98–100]. Notably, ORIGIN-PD analysis revealed that inflammation/immunity module dominance is present in only 3.5% of PD patients. However, it additionally ranked among the top three contributors in MC cases for 35.7% of sporadic patients. This indicates that while peripheral or central immune dysregulation may participate in a subset of patients, it constitutes the primary pathogenic engine in only a minority (38.8% when combining dominance and top-three contributions). Critically, for the remaining 61.2% of patients, inflammation is neither dominant nor a top-tier contributor according to the score. This provides a rational, quantitative basis for enrichment in clinical trials of NLRP3/NF-κB inhibitors: ORIGIN-PD can prospectively exclude both non-responders and minimal responders — two groups that would otherwise contribute to high background noise and attenuated effect sizes — thereby reducing required sample sizes and increasing the probability of detecting a genuine therapeutic signal. Our findings do not diminish the role of neuroinflammation in PD progression, but rather contextualize it as a secondary or amplifying factor in the majority of cases, reinforcing the need for personalized rather than one-size-fits-all immunomodulation.

It should be emphasized that the participant-level module contribution patterns identified by ORIGIN-PD are model-derived representations rather than direct measurements of the true prevalence across patients or the true relative magnitude of pathogenic mechanisms in individual patients. Although each participant can be assigned a ranked contribution profile based on the predefined molecular modules, the observed proportions are influenced by the selected feature set, model coefficients, module definitions, and ranking strategy. Therefore, the frequency with which a given module appears among the dominant or top-ranked contributors should not be interpreted as an exact estimate of the proportion of patients in whom that biological process is truly causal or predominant. Nevertheless, these module contributions are derived from disease-associated changes in feature expression; therefore, for a therapy targeting a given module, patients with a larger contribution of that module would be expected to derive greater benefit than those with a smaller contribution, even though the model-derived contribution cannot be equated with the true magnitude of the underlying pathology. Rather, these module-level distributions should be viewed as an interpretable reference framework for characterizing molecular heterogeneity across patients. Such information may help generate hypotheses regarding biologically distinct patient subsets, support the prioritization of candidate therapeutic pathways, and inform the design of mechanism-oriented clinical studies and drug-development strategies. However, prospective validation using independent cohorts, longitudinal measurements, and orthogonal molecular or functional assays will be required before these contribution patterns can be used for individual-level therapeutic decision-making.

Beyond diagnostic discrimination, the continuous score derived from ORIGIN-PD may offer prognostic or risk-predictive utility. We empirically set a diagnostic threshold of 0.69; in our validation cohorts, healthy controls and most non-PD neurological patients scored below 0.4. This creates an intermediate zone (0.4-0.69) that we hypothesize represents a state of elevated molecular susceptibility, potentially corresponding to prodromal or preclinical PD. This notion is particularly attractive given the current lack of reliable peripheral biomarkers for early risk stratification. However, we acknowledge that this hypothesis requires dedicated longitudinal testing in population-based cohorts with long-term follow-up, as well as cross-validation in diverse ethnic populations (e.g., Asian cohorts) to establish generalizable cutoffs.

Several limitations warrant consideration. First, whole-blood transcriptomics, while minimally invasive[19, 20], is an imperfect surrogate for brain tissue. Our module selection deliberately included blood-specific and brain-mirroring genes, but the absolute contribution scores may not fully recapitulate central pathophysiology. Second, the PPMI cohort, despite rigorous quality control, is predominantly of Caucasian ancestry[101], and validation in independent, ethnically diverse longitudinal cohorts is imperative before clinical deployment. Third, ORIGIN-PD is currently a binary-stage classifier (PD vs. non-PD); its performance in differentiating PD from atypical parkinsonian syndromes (e.g., MSA, PSP) remains unexplored and will require dedicated module recalibration. Lastly, as a cross-sectional model, ORIGIN-PD does not yet capture disease dynamics; future iterations will incorporate serial RNA-seq measurements to model stage-dependent module shifts.

## Conclusion

ORIGIN-PD provides a dual-purpose, interpretable precision framework for stratifying sporadic PD. By assigning each patient a pathogenic-driver profile, it enables (i) rational enrichment for anti-inflammatory clinical trials targeting NLRP3/NF-κB, and (ii) identification of a broader mDA-vulnerable population potentially eligible for emerging WNT-pathway agonists and mDA-protective strategies now advancing through preclinical development. The predominance of the mDA neuron identity/development module, centered on the WNT1/MSX1/LMX1B and NPM1/HSPA8 regulatory network, highlights neurodevelopmental and transcriptional maintenance pathways as major determinants of sporadic PD. ORIGIN-PD therefore offers a rational, mechanism-informed tool for patient selection in targeted trials and may help accelerate the development of precision therapies for sporadic PD.

## Data Availability

All data produced in the present study are available upon reasonable request to the authors

## Abbreviations

PD: Sporadic Parkinson’s disease
mDA: Midbrain dopaminergic
NPM1: Nucleophosmin 1
SNc: Substantia nigra pars compacta
NLRP3: NLR family pyrin domain containing 3
ORIGIN-PD: Octa-module RNA-seq Investigation for Genesis and Initiation of Neurodegenerative Parkinson’s Disease
HC: Healthy control
RIN: RNA integrity number
TMM: Trimmed mean of M-values
log2CPM: log2 counts per million
DEG: Differential-expression gene
LASSO: Least absolute shrinkage and selection operator
PPI: Protein–protein interaction
PPV: Positive predictive value
NPV: Negative predictive value
MC: Multimodule-contribution
MD: Monomodul-dominant
SMDs: Standardized mean differences

## Declarations

### Ethics approval and consent to participate

PPMI is an observational multicenter study in which participants provided written informed consent under protocols approved by the relevant institutional review boards. The present analysis used de-identified PPMI data in accordance with the applicable data-use agreement and PPMI publication requirements.

### Consent for publication

Not applicable.

### Availability of data and materials

Publicly available transcriptomic datasets analyzed in this study were obtained from the Gene Expression Omnibus (GEO), including GSE165082, GSE277709, GSE270454, GSE178491, and GSE185263. Additional datasets, including SoundLife, were accessed and used in accordance with the requirements of their respective data providers or repositories. The source code and supporting files for ORIGIN-PD will be deposited in a public GitHub repository upon publication.

### Authors’ contributions

Conceptualization: YL, CP

Methodology: YL,

Investigation: YL

Visualization: YL, CP

Funding acquisition: CP

Project administration: CP

Supervision: CP

Writing – original draft: YL, CP

Writing – review & editing: YL, XY, CP

## Acknowledgments

We gratefully acknowledge the Parkinson’s Progression Markers Initiative (PPMI) for providing access to the data and resources used in this study. We also sincerely thank all participants who contributed biological samples and clinical information to PPMI, whose participation made this research possible.

---

**Fig. S1.**
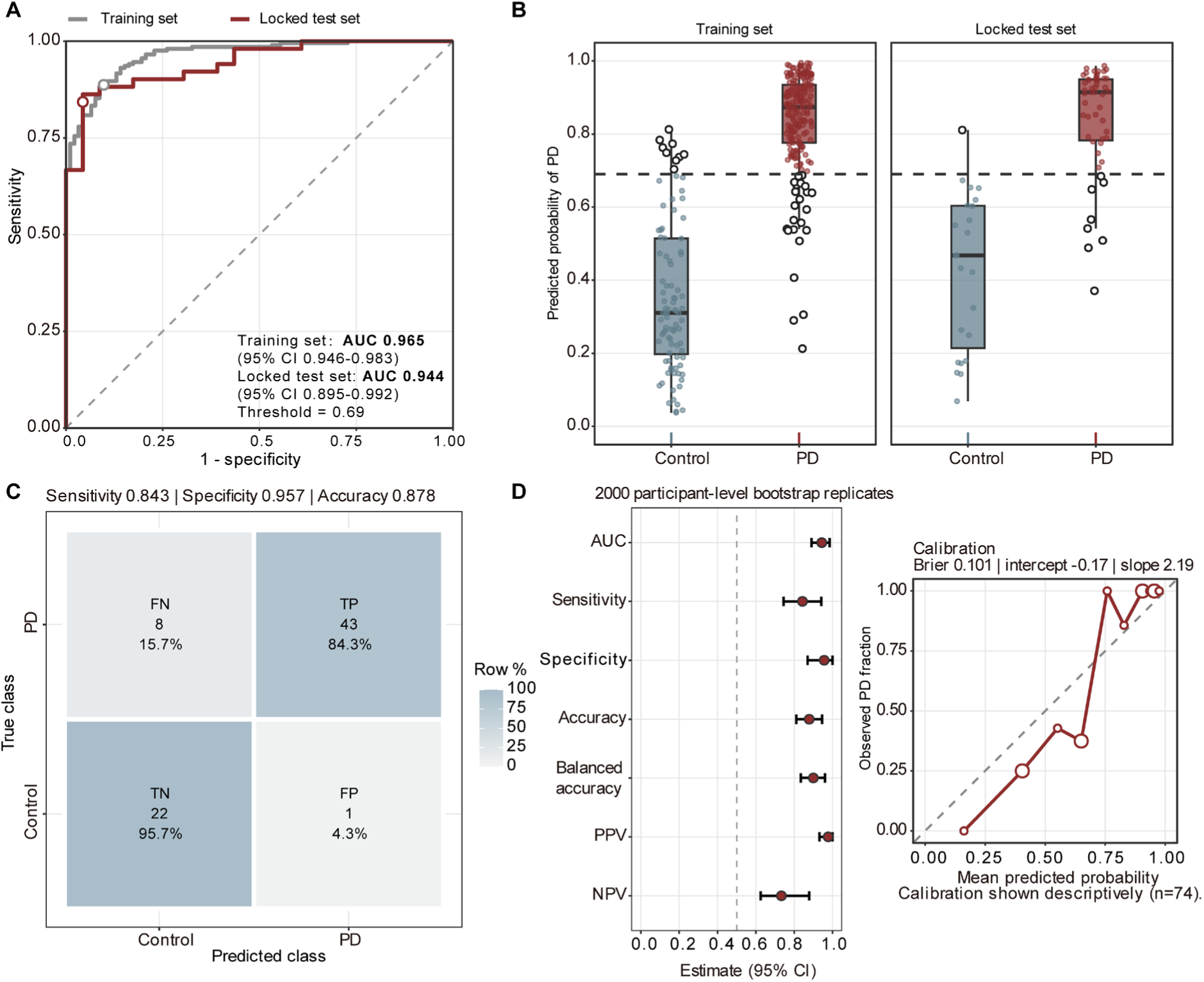
Performance of the Intermediate Model. The intermediate model contained 34 gene features and LowExpr_score before replacement of *HSPA8* by *NPM1*. (**A**) ROC curves in the Training Set (n = 296; 204 PD and 92 HC) and PPMI Locked Test Set (n = 74; 51 PD and 23 HC), with AUCs of 0.965 and 0.944, respectively. Annotated 95% CIs were calculated using the DeLong method; open circles mark the operating points at the 0.69 threshold. (**B**) Assigned PD probabilities by diagnosis and cohort. Points represent participants, open black circles mark classification errors, boxes show medians and interquartile ranges, and whiskers extend to observations within 1.5 times the interquartile range. Dashed horizontal lines mark the threshold. (**C**) Locked Test Set confusion matrix: 43 true positives, 22 true negatives, 1 false positive and 8 false negatives, corresponding to sensitivity 0.843 and specificity 0.957. Cell percentages are calculated within true-class rows. (**D**) Locked Test Set ROC-AUC, sensitivity, specificity, accuracy, balanced accuracy, positive predictive value and negative predictive value with 95% percentile CIs from 2,000 outcome-stratified bootstrap replicates (left), and quantile-binned calibration (right). Calibration-point size indicates bin size; the diagonal represents ideal calibration. The Brier score was 0.101, calibration-in-the-large was −0.17, and calibration slope was 2.19.

**Fig. S2.**
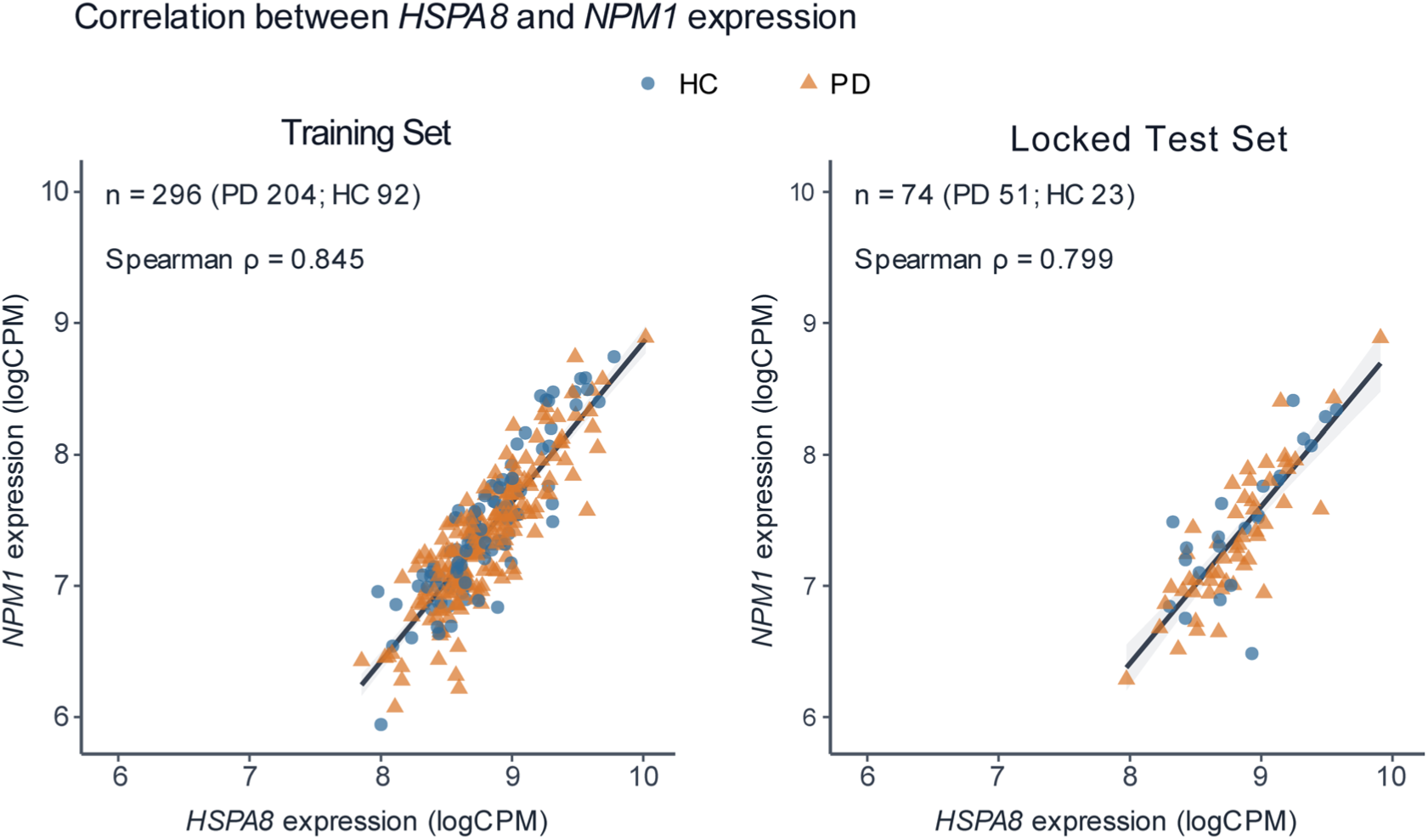
Correlation between *HSPA8* and *NPM1* expression in the PPMI Training Set and Locked Test Set. *HSPA8* and *NPM1* expression in the Training Set (left; n = 296) and PPMI Locked Test Set (right; n = 74). Each point represents one participant; blue circles indicate HC and orange triangles indicate PD. Expression is shown on the log2 counts-per-million scale generated by the formal frozen-reference normalization workflow. Black fitted lines and grey 95% confidence bands summarize the pooled PD and HC observations within each cohort. The displayed Spearman correlations were 0.845 in the Training Set and 0.799 in the Locked Test Set. These are descriptive within-cohort associations across diagnoses, not diagnosis-adjusted correlations, evidence of biological interchangeability, or causal evidence for the *HSPA8*-to-*NPM1* model refinement. HC, healthy control; PD, Parkinson’s disease; PPMI, Parkinson’s Progression Markers Initiative.

**Fig. S3.**
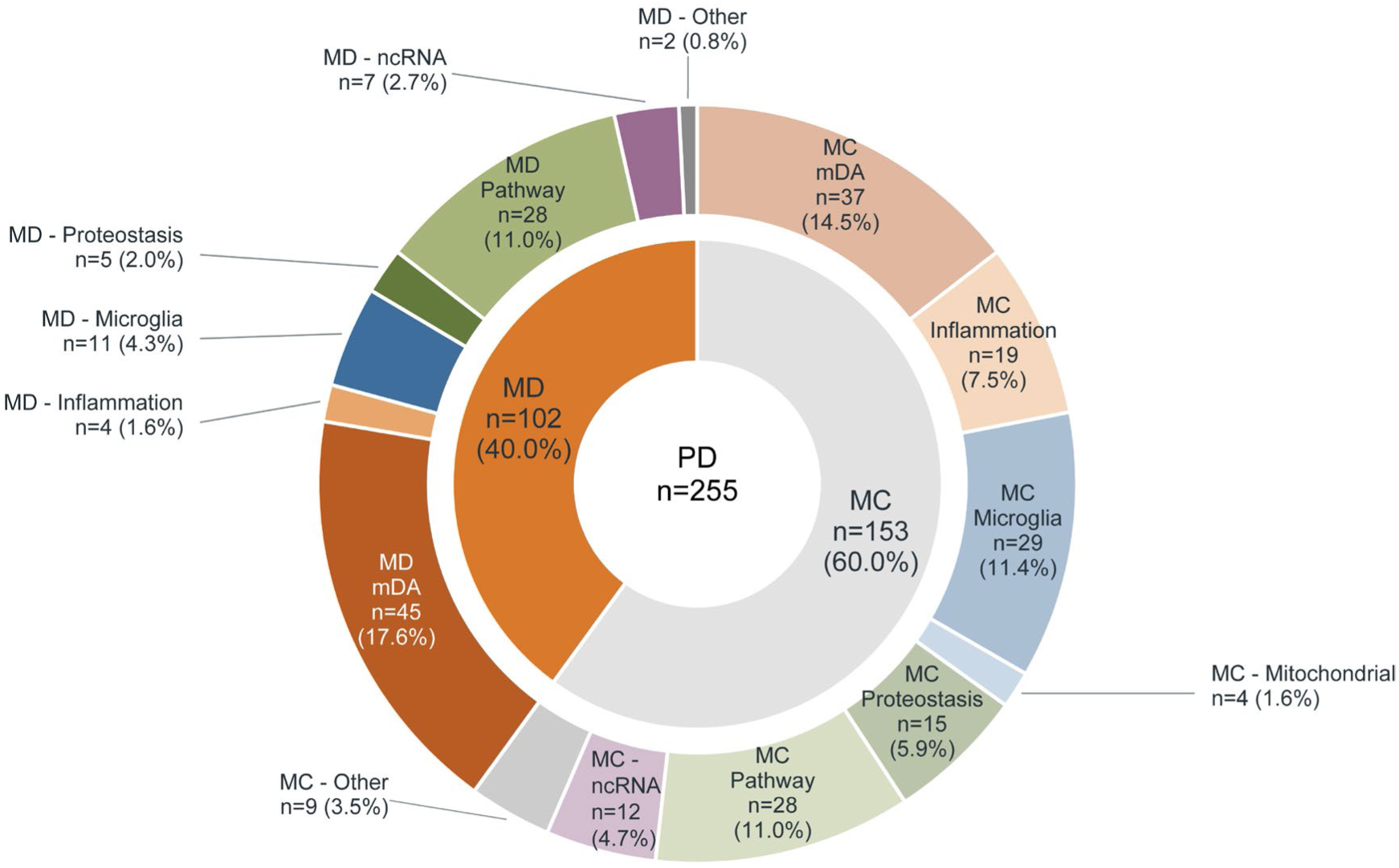
Distribution of functional-module contribution patterns in the Intermediate Model among participants with PD. The donut chart summarizes participant-level molecular contribution patterns of the Intermediate Model across all 255 participants with PD. Gene-level predictors were assigned a priori to eight functional modules: mDA neuron identity, inflammation/immunity, oxidative/mitochondrial, non-coding RNA, proteostasis/clearance, pathway-related, microglia/glial, and other. The proportions shown represent the percentage of PD participants assigned to each indicated contribution pattern. LowExpr_score was retained as an independent composite predictor and was not included in the eight-module ranking.

**Table S1.**
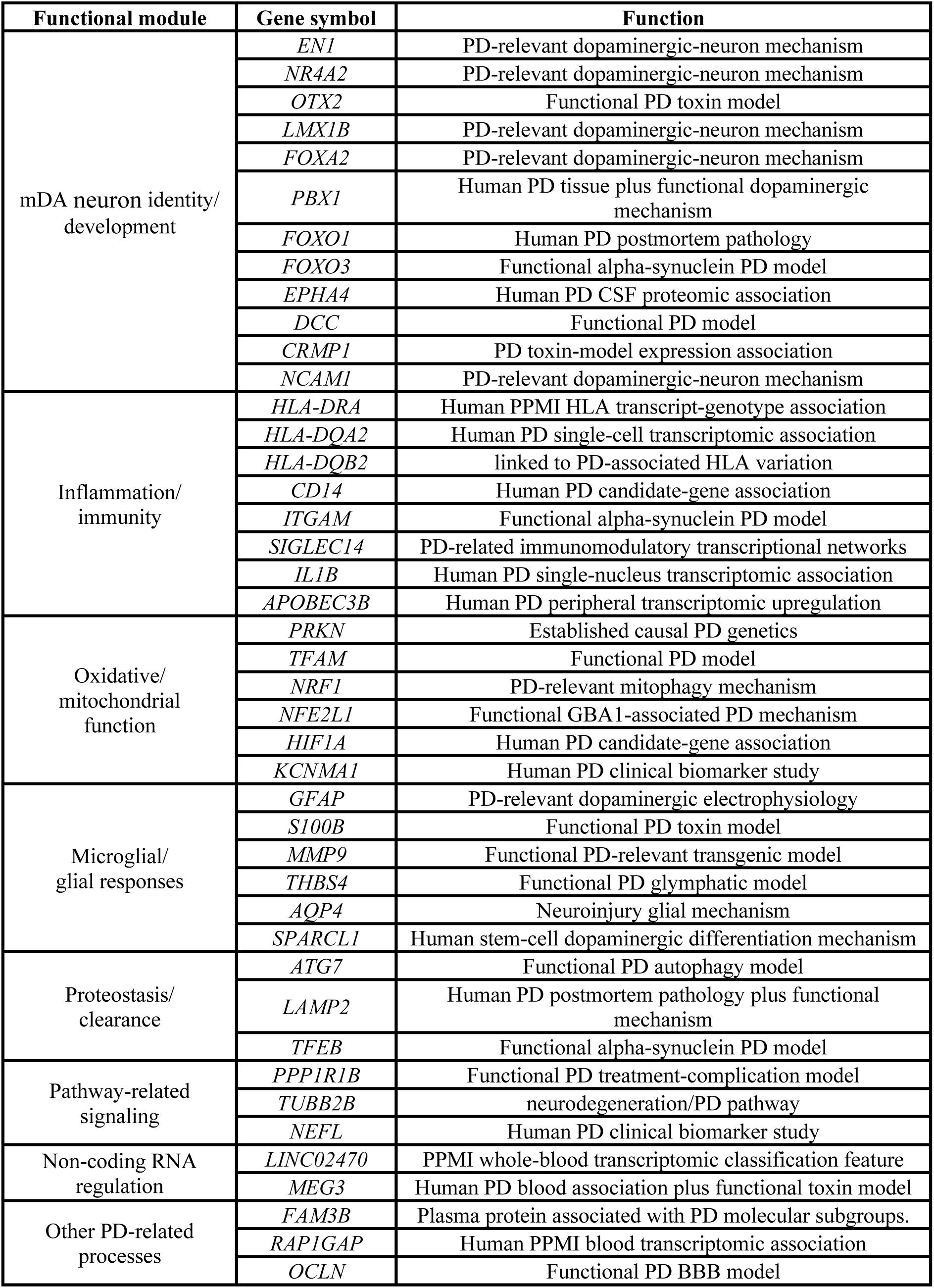
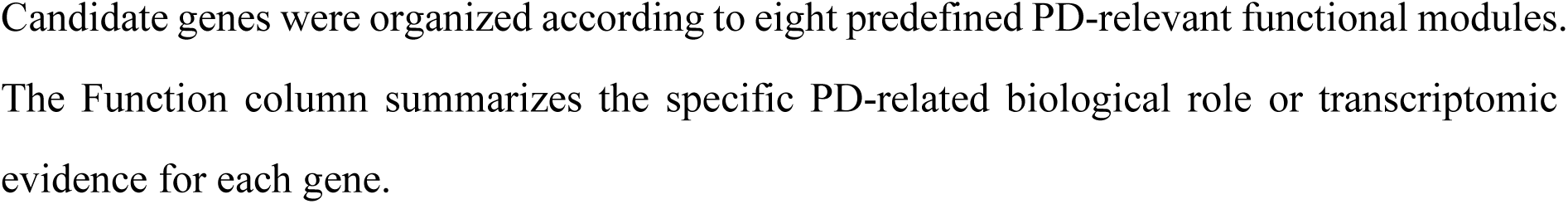
Prespecified candidate genes across the eight PD-related functional modules.

| Functional module | Gene symbol | Function |
| --- | --- | --- |
| mDA neuron identity/<br>development | *EN1* | PD-relevant dopaminergic-neuron mechanism |
|  | *NR4A2* | PD-relevant dopaminergic-neuron mechanism |
|  | *OTX2* | Functional PD toxin model |
|  | *LMX1B* | PD-relevant dopaminergic-neuron mechanism |
|  | *FOXA2* | PD-relevant dopaminergic-neuron mechanism |
|  | *PBX1* | Human PD tissue plus functional dopaminergic mechanism |
|  | *FOXO1* | Human PD postmortem pathology |
|  | *FOXO3* | Functional alpha-synuclein PD model |
|  | *EPHA4* | Human PD CSF proteomic association |
|  | *DCC* | Functional PD model |
|  | *CRMP1* | PD toxin-model expression association |
|  | *NCAM1* | PD-relevant dopaminergic-neuron mechanism |
| Inflammation/<br>immunity | *HLA-DRA* | Human PPMI HLA transcript-genotype association |
|  | *HLA-DQA2* | Human PD single-cell transcriptomic association |
|  | *HLA-DQB2* | linked to PD-associated HLA variation |
|  | *CD14* | Human PD candidate-gene association |
|  | *ITGAM* | Functional alpha-synuclein PD model |
|  | *SIGLEC14* | PD-related immunomodulatory transcriptional networks |
|  | *IL1B* | Human PD single-nucleus transcriptomic association |
|  | *APOBEC3B* | Human PD peripheral transcriptomic upregulation |
| Oxidative/<br>mitochondrial<br>function | *PRKN* | Established causal PD genetics |
|  | *TFAM* | Functional PD model |
|  | *NRF1* | PD-relevant mitophagy mechanism |
|  | *NFE2L1* | Functional GBA1-associated PD mechanism |
|  | *HIF1A* | Human PD candidate-gene association |
|  | *KCNMA1* | Human PD clinical biomarker study |
| Microglial/<br>glial responses | *GFAP* | PD-relevant dopaminergic electrophysiology |
|  | *S100B* | Functional PD toxin model |
|  | *MMP9* | Functional PD-relevant transgenic model |
|  | *THBS4* | Functional PD glymphatic model |
|  | *AQP4* | Neuroinjury glial mechanism |
|  | *SPARCL1* | Human stem-cell dopaminergic differentiation mechanism |
| Proteostasis/<br>clearance | *ATG7* | Functional PD autophagy model |
|  | *LAMP2* | Human PD postmortem pathology plus functional mechanism |
|  | *TFEB* | Functional alpha-synuclein PD model |
| Pathway-related<br>signaling | *PPP1R1B* | Functional PD treatment-complication model |
|  | *TUBB2B* | neurodegeneration/PD pathway |
|  | *NEFL* | Human PD clinical biomarker study |
| Non-coding RNA<br>regulation | *LINC02470* | PPMI whole-blood transcriptomic classification feature |
|  | *MEG3* | Human PD blood association plus functional toxin model |
| Other PD-related<br>processes | *FAM3B* | Plasma protein associated with PD molecular subgroups. |
|  | *RAP1GAP* | Human PPMI blood transcriptomic association |
|  | *OCLN* | Functional PD BBB model |

Candidate genes were organized according to eight predefined PD-relevant functional modules. The Function column summarizes the specific PD-related biological role or transcriptomic evidence for each gene.

**Table S2.**
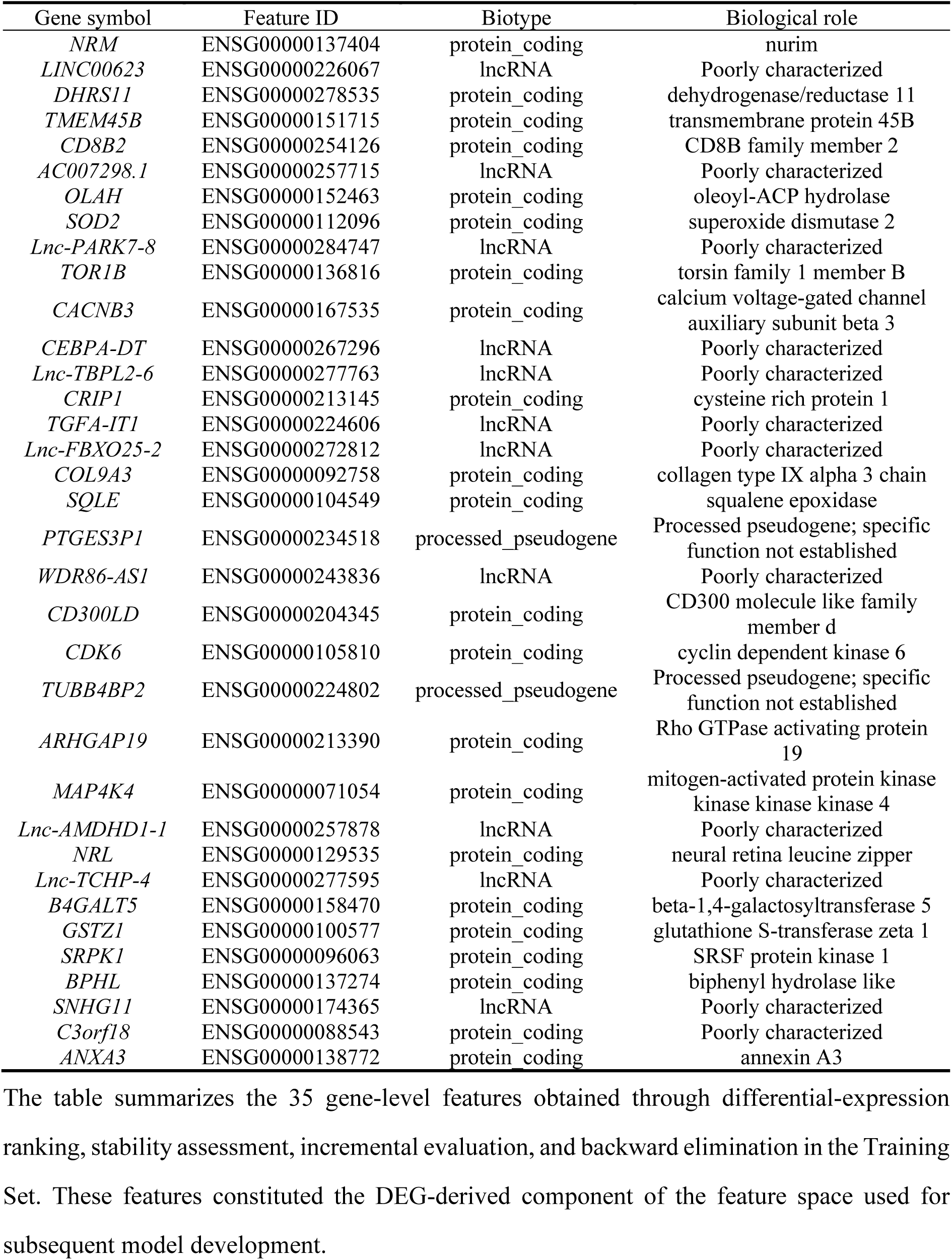
DEG-derived gene features retained before development of the Primary Model.

**Table S3.**
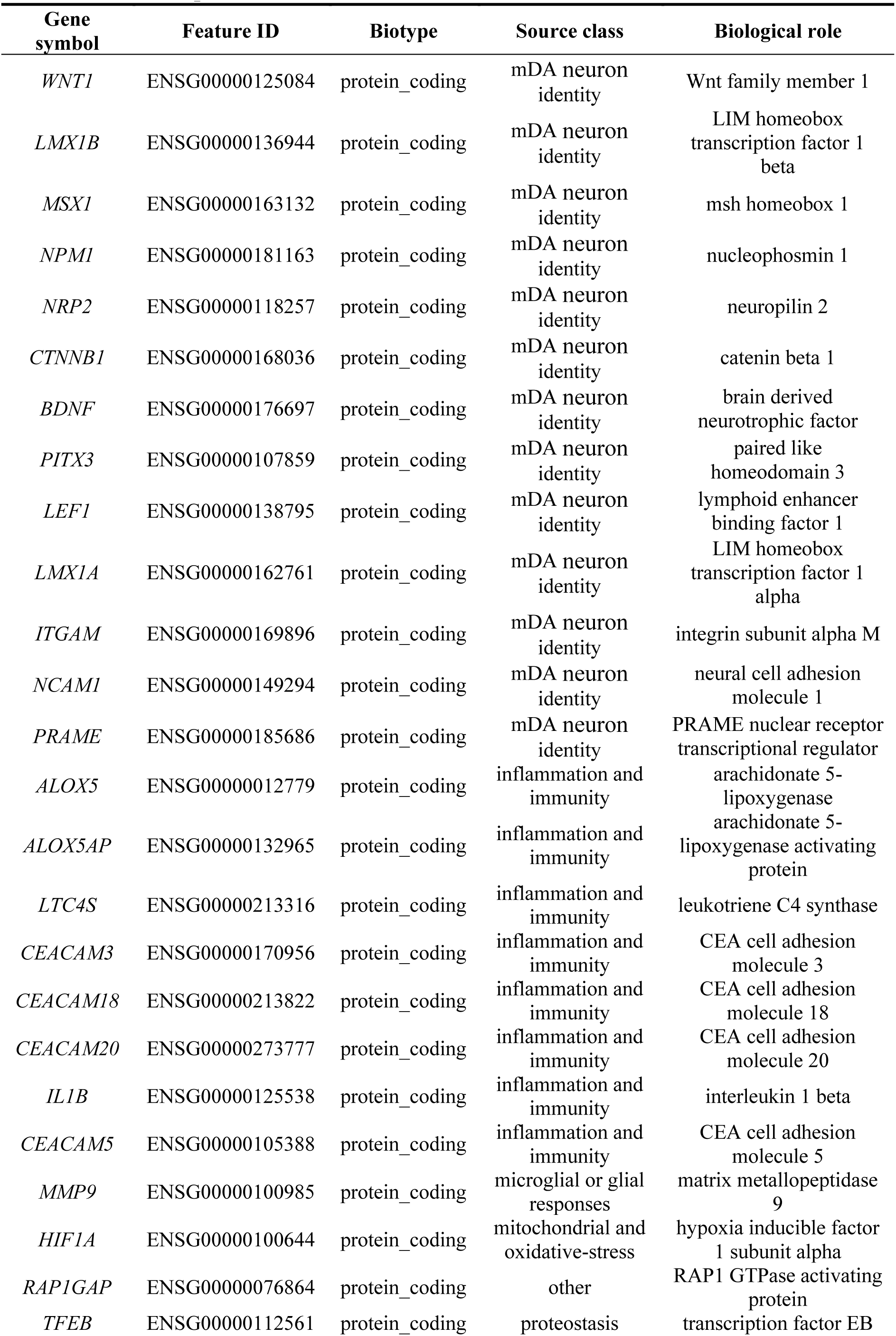
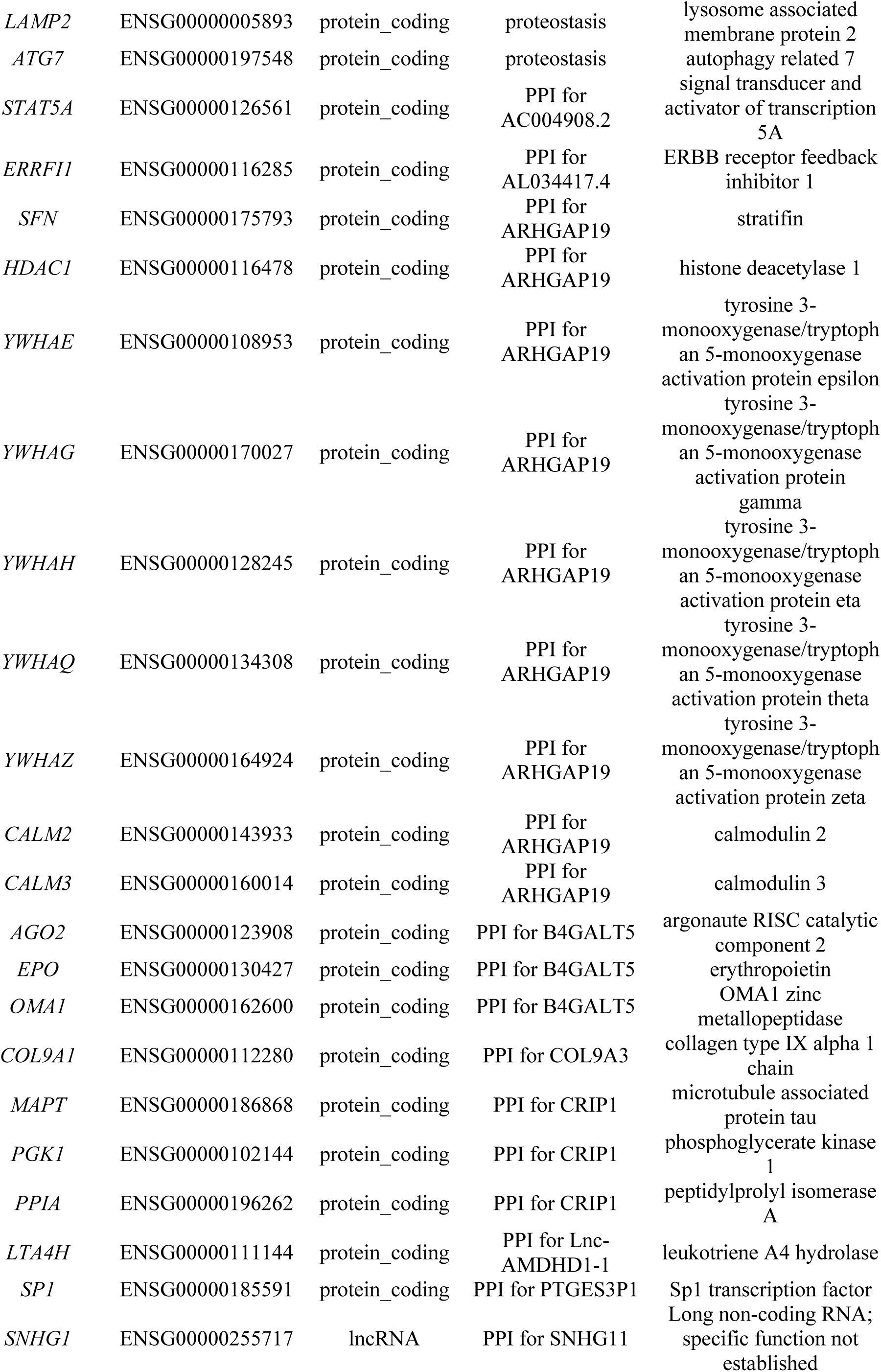
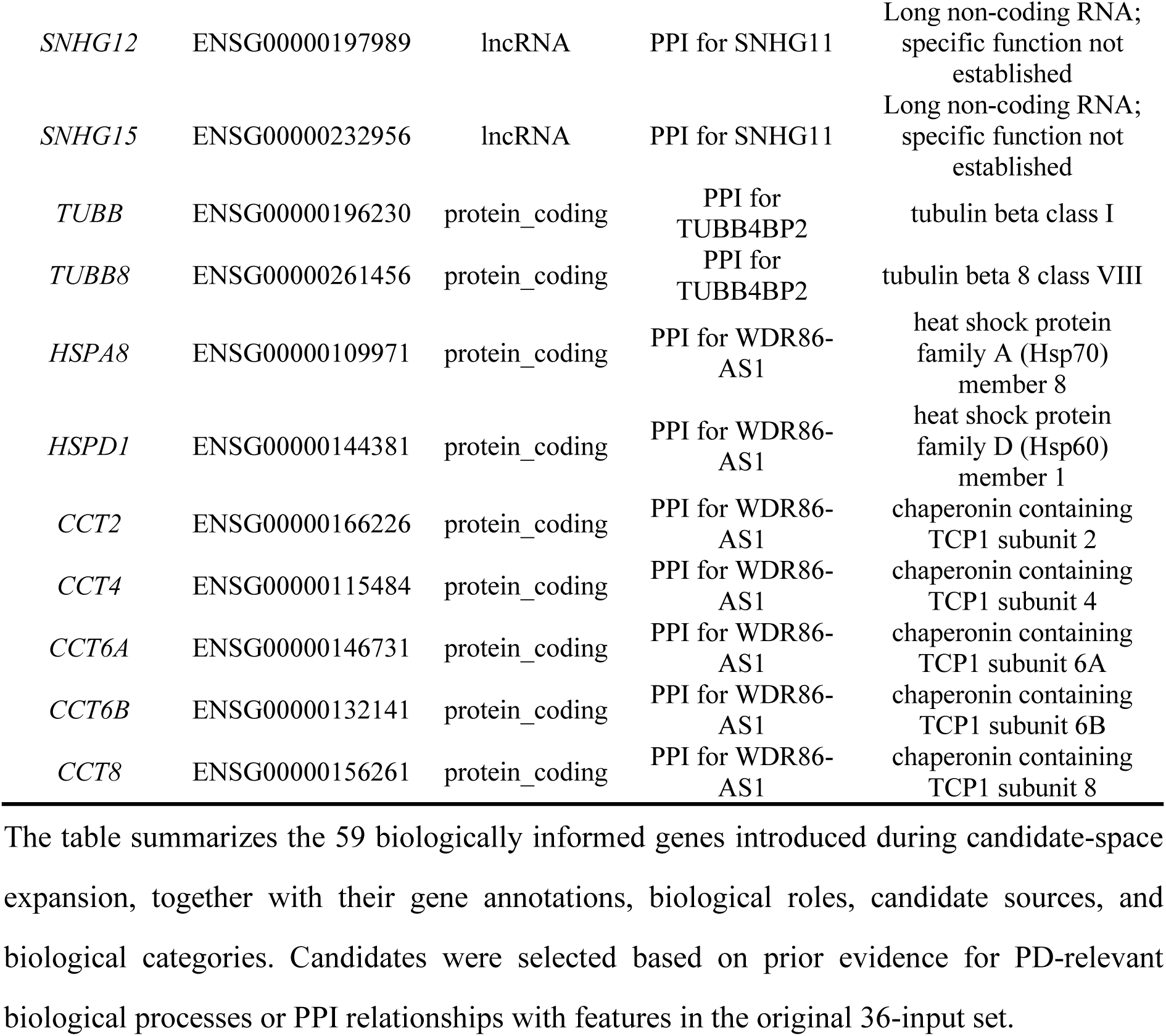
Biologically informed genes introduced during expansion of the ORIGIN-PD candidate feature space.

**Table S4.** Comparison of participant-level module rankings between ORIGIN-PD and the Intermediate Model.

| Functional module | Origin-PD |  | Intermediate Model |  |
| --- | --- | --- | --- | --- |
|  | Rank1–5 | Top5 | Rank1–5 | Top5 |
| mDA neuron identity/development | 35.7% / 20.4% / 8.2% / 10.6% / 7.5% | 82.4% | 32.2% / 15.7% / 10.6% / 10.2% / 9.4% | 78.0% |
| Inflammation/immunity | 9.4% / 11.8% / 17.6% / 16.5% / 15.7% | 71.0% | 9.0% / 12.5% / 12.9% / 15.3% / 12.5% | 62.4% |
| Microglia/glia | 18.4% / 25.1% / 16.5% / 10.2% / 8.2% | 78.4% | 15.7% / 23.1% / 18.0% / 8.2% / 8.2% | 73.3% |
| Oxidative/mitochondrial | 2.0% / 3.9% / 11.0% / 14.5% / 17.3% | 48.6% | 1.6% / 5.5% / 7.1% / 9.8% / 14.1% | 38.0% |
| Proteostasis/clearance | 0.0% / 0.0% / 0.0% / 2.0% / 4.3% | 6.3% | 7.8% / 9.4% / 15.3% / 15.3% / 15.7% | 63.5% |
| Pathway-related | 22.0% / 17.6% / 20.0% / 16.1% / 9.0% | 84.7% | 22.0% / 14.5% / 18.0% / 14.1% / 9.8% | 78.4% |
| Non-coding RNA | 8.2% / 12.5% / 17.3% / 17.3% / 17.3% | 72.5% | 7.5% / 13.3% / 9.8% / 16.9% / 14.5% | 62.0% |
| Other | 4.3% / 8.6% / 9.4% / 12.9% / 20.8% | 56.1% | 4.3% / 5.9% / 8.2% / 10.2% / 15.7% | 44.3% |

The table summarizes the distribution of the eight predefined biological modules across ranks 1–5 among 255 participants with PD. Module rankings were determined for each participant according to the absolute magnitude of the signed module contribution.

